# Neuron-derived circulating miRNAs reveal lead (Pb) as a key component of metal mixtures exposure

**DOI:** 10.1101/2025.02.10.25322027

**Authors:** Janina Manzieri Prado-Rico, Alejandra Bargues-Carot, Yuka Imamura Kawasawa, Jiazhang Cai, Jeff D. Yanosky, Lan Kong, Gary Zenitsky, Huajun Jin, Andre L. Garao Rico, Molly A. Hall, Mechelle M. Lewis, Ping Ma, Vellareddy Anantharam, Arthi Kanthasamy, Richard B. Mailman, Anumantha G. Kanthasamy, Xuemei Huang

## Abstract

Environmental exposures rarely occur in isolation, yet biomarkers capturing early brain responses to complex metal mixtures remain limited. Neuron-derived miRNAs detectable in peripheral blood may provide minimally invasive indicators of brain-related molecular processes. We profiled miRNAs from neuron-derived extracellular vesicles in 66 adults and identified 50 dysregulated miRNAs. Among these, miR-16-5p, miR-93-5p, and miR-486-5p were reduced in individuals with higher exposure levels. Metal mixture models identified Pb as the metal most consistently associated with these miRNAs. To explore the translational relevance of these findings, we integrated brain MRI measures and observed that mediation analyses suggested miR-16-5p may represent a potential pathway linking Pb exposure to iron-sensitive MRI signals (R2*, a marker of brain iron) in the red nucleus. Together, these results suggest circulating neuron-derived miRNAs may capture molecular signatures linking complex metal mixtures, with Pb as a key component, to neuronal regulatory pathways and early brain-related perturbation to real-world exposures.

## Introduction

Humans are chronically exposed to complex mixtures of environmental metals through air pollution, occupational and recreational activities, contaminated soil, food, and water,^1,2^ representing a widespread public health concern. These mixtures include both essential elements (e.g., manganese [Mn], iron [Fe], copper [Cu]) and toxicants (e.g., lead [Pb]), which can induce oxidative stress, impair mitochondrial function, and promote inflammatory responses,^3–6^ mechanisms implicated in neurodegenerative diseases (NDDs).^7–10^ Understanding how these metal mixtures exposures influence molecular pathways relevant to brain health remains an important question in environmental health research.

MicroRNAs (miRNAs) are small non-coding RNAs that regulate gene expression and play key roles cellular homeostasis, including pathways related to oxidative stress, inflammation, and neurodegeneration.^11–14^ Circulating miRNAs can be detected in plasma, serum, and cerebrospinal fluid (CSF),^15^ where they are stabilized by binding to RNA-binding proteins^16^ or lipoproteins^17^ and by being encapsulated within extracellular vesicles (EVs).^18^ Notably, EVs can cross the blood-brain barrier (BBB), allowing central nervous system (CNS)-derived miRNAs to enter the peripheral circulation.^19,20^ Because miRNAs regulate gene expression at the post-transcriptional level, changes in their circulating levels may reflect perturbations in gene regulatory networks associated with environmental exposures.

An emerging view in environmental health research suggests that blood-based molecular signatures, including regulatory RNAs, may reveal early biological responses to metal mixtures exposure before the clinical onset of diseases.^21,22^ Peripheral biomarkers capable of reflecting brain-related biological processes are limited, thus circulating neuron-derived EVs miRNAs have emerged as promising minimally invasive biomarkers for monitoring CNS-related molecular processes, potentially reflecting early neurobiological stress and neurodegeneration.^15,19^

Growing evidence indicates that exposure to metal-rich particulate and other environmental toxicants can alter miRNA expression.^12,23–26^ Importantly, environmental exposures rarely occur in isolation, as individuals are typically exposed to complex metal mixtures with potential additive or synergistic effects.^12,27^ Although miRNA dysregulation has been associated with both environmental toxicants and pathways involved in NDDs,^7–10,28^ it remains unclear whether circulating neuron-derived EVs miRNAs reflect the neurobiological impact of metal mixtures.

To address this gap, we profiled neuron-derived miRNAs isolated from EVs in a clinically asymptomatic cohort with heterogeneous metal exposures. We integrated differential expression analyses, exposure–miRNA correlation analyses, and Bayesian kernel machine regression (BKMR) to evaluate exposure-response relationship between circulating miRNAs and metal mixture exposure. By combining neuron-derived molecular markers with mixture modeling, this study connects circulating miRNAs detectable in peripheral blood to brain-related molecular perturbation associated with metal mixtures exposure. To explore the potential translational relevance of linking real-world exposure, molecular responses, and brain imaging signals (Figure 1), we evaluated whether circulating miRNAs associated with metal exposure were related to iron-sensitive MRI markers (R2*).

**Figure 1.**
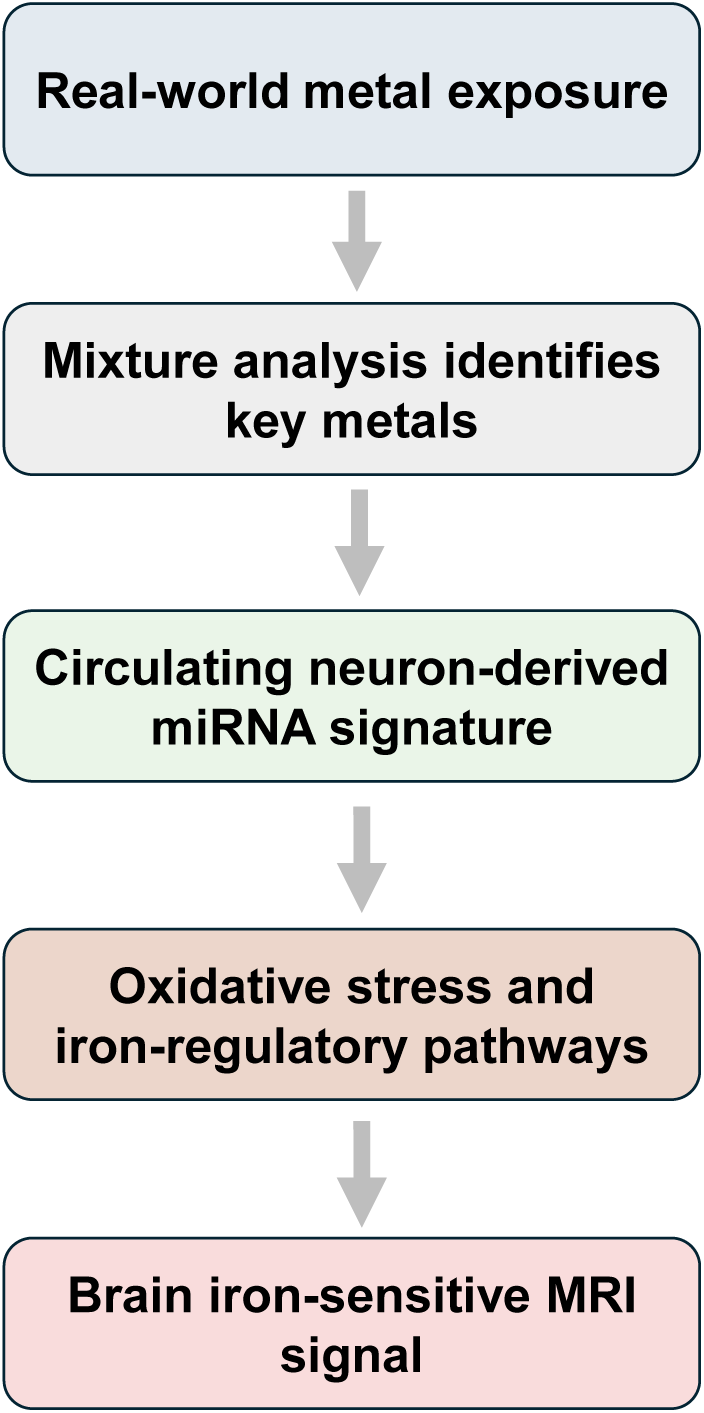
Conceptual framework of the translational approach used in this study. Real-world metal exposure was examined using mixture models to identify metals associated with circulating neuron-derived miRNAs. These miRNAs may capture neuronal molecular responses related to metals exposure, which may in turn relate to iron-sensitive MRI signals in the brain.

## Methods

### Subjects

Demographic data and miRNA measures were obtained from a subset of 66 participants who were part of a larger group of participants enrolled in previous studies.^29,30^ Participants were categorized as non-exposed (n = 27) or exposed (n = 39), based on their lifetime exposure to welding fumes, allowing the evaluation of exposure–response patterns across a gradient of exposure. Exposed participants were classified as individuals who had engaged in welding activities at any point in their lives, whereas non-exposed participants included individuals in various occupations with no history of welding. These participants were recruited from labor unions and surrounding communities adjacent to Hershey, Pennsylvania, USA.

All participants were male reflecting the overall demographics of this occupation in central Pennsylvania. They were free of neurological disorders or motor dysfunction at the time of study, as assessed by the Movement Disorders Society Unified Parkinson’s Disease Rating Scale-motor score (MDS-UPDRS-III) with a threshold score <15.^30^ The protocol was reviewed and approved by the Penn State College of Medicine Institutional Review Board. Written informed consent was obtained from all participants, and the study was conducted consistent with the Declaration of Helsinki.

### Blood collection and serum preparation

For this study, blood samples were collected from participants following an 8-12-hour overnight fast. Whole-blood metal concentrations were measured using previously described protocols.^29^ Within 30–60 min after the blood draw, samples were centrifuged at 1,500 x g for 15 min at 4°C. One-mL aliquots of the supernatant were pipetted into cryovials on ice and then stored at −80°C. For exosome and microvesicle analyses, serum samples were thawed, and 100 μL was pipetted into a separate cryovial tube and used as described below. Serum was chosen over plasma to reduce the background levels of proteins, lipids, and sugars that may interfere with the isolation and characterization of EVs, ensuring higher purity and assay sensitivity.^31^

### Extracellular vesicle and miRNA isolation

The EVs, including exosomes and microvesicles, were purified from the samples to separate free circulating miRNAs from those contained within the vesicles of interest.^32^ ExoQuick Exosome Precipitation Solution kit (Systems Biosciences) was used to extract EVs from the samples. Serum (100 μL) was centrifuged at 3,000 x g for 15 min at room temperature to remove cells and cellular debris. The supernatant was transferred to a sterile vessel, 25.2 μL of ExoQuick Exosome Precipitation Solution was added, refrigerated at 4°C for 30 min, and centrifuged at 1,500 x g for 30 min at 4°C. The supernatant was aspirated, leaving the EVs as a white pellet. Another centrifugation at 1,500 x g for 5 min was done to remove traces of ExoQuick by aspiration. The EV pellet was resuspended in 100 μL phosphate-buffered saline (137 mM NaCl, 2.7 mM KCl, 10 mM Na_2_HPO_4_, and 1.8 mM KH_2_PO_4_).

The resuspended pellets were analyzed to confirm that the purified EVs had the expected size and concentration measurements (see details in ^31^). Briefly, transmission electron microscopy was used to confirm that the exosomes in the buffer had an expected size of 40–200 nm^33^ and microvesicles ranging from 200–1,000 nm. EVs with neuronal origins were extracted from the solution by capturing exosomes and microvesicles expressing the neuron-specific marker CD171 (L1 cell adhesion molecule).^19^ A biotinylated anti-human CD171 antibody (eBio5G3, Affymetrix) was used to bind vesicles expressing CD171. Then, streptavidin-conjugated magnetic beads (10608D, ThermoFisher Scientific) were added to the solution to bind to the CD171 antibody. Neuronal EVs bound to anti-CD171 and streptavidin beads were pulled down magnetically. The captured neuron-derived EVs were lysed with IGEPAL CA-630 (Sigma-Aldrich) to free the miRNAs into solution; IGEPAL was added to 1% of the final concentration. The miRNAs derived from these neuronal EVs were used for sequencing library preparation.

### MiRNA-sequencing (RNA-seq)

The miRNA sequencing libraries were generated from serum-derived EV miRNAs using the CleanTag Small RNA Library Prep Kit (TriLink Biotechnologies) for downstream miRNA expression analysis. Individually barcoded libraries were mixed equimolarly and subjected to sequencing with an Illumina NovaSeq 6000.

### Welding-related exposure assessment

Welding-related exposure was assessed by a work history (WH)^29,30^ questionnaires that collected job information over the individual’s working lifetime, emphasizing welding and other jobs associated with welding-related metal fume exposure. Responses allowed us to calculate cumulative lifetime years spent welding (YrsW), and an estimate of lifetime cumulative exposure to metal fumes during welding [ELT (units of mg*years*m^−3^)],^29,30^ which integrated welding tasks type (*e.g.*, brazing, soldering, *etc.*), time spent performing each activity, and the use of local ventilation.^29,30^

An additional supplementary exposure questionnaire (SEQ)^29,30^ focused on the 90-day period prior to the study visit and the time spent welding, types of metal welded, and type of welding performed. The exposure metrics derived from the SEQ included hours of welding (HrsW) and an estimate of recent exposure in the 90-day period preceding the study visit (E90).^29,30^

### Data processing and statistical analysis

Demographic and clinical data were compared across exposure groups (non-exposed vs. exposed) using one-way analysis of variance (ANOVA), chi-square, or Mann-Whitney U tests, as appropriate. Blood metal concentrations were evaluated for differences across exposure groups using analysis of covariance (ANCOVA), adjusting for age.

To examine individual metal contributions and effects of co-exposure to metal mixtures on miRNA expression, we applied a multi-step analytical approach:

1. *<u>Differential expression (DE) analysis based on exposure status:</u>* DE analyses were performed between exposure groups to identify candidate miRNAs associated with higher exposure contrast. The deep-sequencing data were subjected to a series of steps from preprocessing to generating an MA plot, volcano plot, hierarchical clustering heatmap, principal coordinate analysis (PCoA) plot, and linear discriminant analysis (LDA) effect size (LEfSe) plot. The read number of each miRNA was used for deep-sequencing analysis. Any miRNAs with missing records were removed from the miRNA count matrix. Subsequently, only miRNAs with read counts > 10 in more than three samples were kept. The count matrix was then converted to a DESeq object for further analysis using the DESeq2 package (version 1.40.2) in R (version 4.3.1). The DESeq2 summarizes the mean for each miRNA and calculates the log_2_-fold change of each miRNA between exposure groups based on the application of an empirical Bayes shrinkage procedure to the coefficients of a negative binomial regression model.^34^ Based on this method, *p*-values and related statistics were obtained to test the significance of the differential expression of each miRNA.
2. *<u>Exposure–miRNA association analyses:</u> Pearson* partial correlations were conducted across the full cohort to evaluate associations between miRNAs expression and whole-blood metal concentrations [copper (Cu), iron (Fe), potassium (K), magnesium (Mg), manganese (Mn), sodium (Na), lead (Pb), selenium (Se), strontium (Sr), and zinc (Zn)] both natural log-transformed, across de exposure gradient. We first adjusted for age and subsequently for both age and exposure groups to evaluate the robustness of associations. miRNAs that were significantly correlated with whole-blood metal concentration levels were further investigated in the scientific literature to determine if they had been linked to metals exposure or other toxicants, neuroinflammation, Alzheimer’s disease (AD) and related dementias (ADRD), Parkinson’s disease (PD), and other NDDs. Associations between differentially expressed miRNAs and long-term (YrsW, ELT) and short-term (HrsW, E90) welding exposure measures were also examined using *Pearson* partial correlations, first adjusted for age and subsequently for both age and exposure group.
3. *<u>Metal mixture modeling:</u>* To account for potential nonlinear exposure–response relationships, interactions among metals, and joint mixture effects, we applied BKMR. We modeled the relationships between the 10-metal mixture and miRNA expression across the cohort. Models were run for 10,000 iterations using the Markov chain Monte Carlo algorithm in R (v3.6.1) with the BKMR package.^35^ Posterior inclusion probabilities (PIPs), overall mixture effects, single and bivariate exposure–response relationships, and single-variable risk were estimated. Because exposure group was defined to increase exposure contrast within the cohort, primary analyses evaluated exposure–response relationships across the full exposure gradient adjusting for age. Additional models were further adjusted for age and exposure groups to assess the robustness of the findings.
4. *<u>Brain MRI susceptibility measure (R2*) and exploratory mediation analysis:</u>* R2* metrics were used as an iron-sensitive MRI susceptibility measure that can reflect alterations in brain iron homeostasis associated with metal-induced oxidative stress.^4,36^ R2* values were obtained from predefined regions of interest (ROI) in participants from the same cohort, as previously described.^4^ *Pearson* partial correlation adjusted for age were first used to examine associations between circulating miRNA expression and R2* values across ROIs. For ROIs showing statistically significant correlations, exploratory mediation analyses were subsequently conducted using linear regression with bootstrap resampling (5,000 iterations) to estimate indirect effects and corresponding confidence intervals. Exposure (X) was whole-blood metal, mediators (M) individual miRNA expression levels, and the outcome (Y) R2* values from ROIs.^4^ Models were adjusted for age and subsequently for age and exposure group to assess robustness.

Uncorrected p-values are reported to examine associations. To account for multiple comparisons, p-values were additionally adjusted using the Benjamini–Hochberg false discovery rate (FDR), The results that remained significant after FDR correction are indicated. Given the exploratory nature of this study, primary inferences were based on consistency of associations across analytical approaches, with FDR-adjusted p-values used to assess robustness. Statistical significance was defined as α = 0.05. All statistical analyses were conducted using SPSS version 27.0 and Python.

## Results

### Demographic and clinical data

Demographic characteristics are summarized in Table 1. Participants did not differ significantly in age *[F(1,65) = 1.70, p-value = 0.196]*. Participants classified as non-exposed had a significantly higher level of education compared to exposed participants *[F(1,65) = 48.85, p-value < 0.001]*, whereas exposed participants had a significantly higher body mass index (BMI) *[F(1,64) = 7.02, p-value = 0.010]* (Table 1). Non-exposed and exposed participants did not differ significantly in UPDRS-III subscore *[U = 499.0, Z = −0.37, p = 0.711]*.

**Table 1.** Demographic, clinical, welding-related exposure measures, and whole-blood metals concentration levels for non-exposed (n=27) and exposed (n=39) participants.

|  | Non-exposed (n=27) |  |  |  | Exposed (n=39) |  |  |  |  |
| --- | --- | --- | --- | --- | --- | --- | --- | --- | --- |
| | Min | Mean (SD) | Median (Q1 - Q3) | Max | Min | Mean ± SD | Median (Q1 - Q3) | Max | $\eta_p^2$; p-value |
| **<u>Demographic and clinical data</u>** |  |  |  |  |  |  |  |  |  |
| Age (years) | 28 | 44 ± 10 | 42 (38 – 53) | 61 | 24 | 47 ± 10 | 51 (40 – 55) | 65 | 0.026; 0.196 |
| Education (years) | 12 | 16 ± 2 | 16 (16 – 18) | 22 | 11 | 13 ± 1 | 12 (12 – 14) | 16 | **0.950; <0.001** |
| BMI (kg/m<sup>2</sup>) | 21.0 | 26.3 ± 3.4 | 26.6 (23.1 – 29.2) | 32.9 | 21.1 | 29.0 ± 4.3 | 28.5 (26.1 – 30.9) | 40.7 | **0.100; 0.010** |
| UPDRS-III (motor scores) | 0 | 1 ± 2 | 1 (0 – 2) | 8 | 0 | 2 ± 2 | 1 (0 – 2) | 7 | 0.711 |
| Ever-smokers (n, %)* | ---- | 7 (26%) | ---- | ---- | ---- | 24 (61%) | ---- | ---- | **0.006** |
| Pack-years smoking | 0.2 | 23.0 ± 29.6 | 16.5 (0.8 – 38) | 80.0 | 0.5 | 19.8 ± 21.3 | 12.7 (7.4 – 28.3) | 97.5 | 0.911 |
| Any alcohol consumption n, %* | ---- | 12 (44%) | ---- | ---- | ---- | 20 (51%) | ---- | ---- | 0.624 |
| Alcohol consumption (drinks/week) | 2 | 6 ± 6 | 5 (2 – 6) | 20 | 1 | 5 ± 3 | 5 (3 – 6) | 16 | 0.557 |
| **<u>Whole-blood metal concentration levels</u>** |  |  |  |  |  |  |  |  |  |
| Whole blood Cu (μg/L) | 502 | 729 ± 132 | 742 (610 – 823) | 957 | 553 | 900 ± 125 | 910 (827 – 996) | 1,111 | **0.361; <0.001** |
| Whole blood Fe (mg/L) | 198 | 489 ± 79 | 504 (474 – 540) | 583 | 397 | 556 ± 55 | 558 (533 – 593) | 673 | **0.172; 0.003** |
| Whole blood K (mg/L) | 1100 | 1,736 ± 316 | 1,650 (1540 – 2018) | 2,300 | 1,290 | 2,163 ± 364 | 2,197 (1833 – 2460) | 2,725 | **0.389; <0.001** |
| Whole blood Mg (mg/L)<sup>+</sup> | 22.3 | 35.1 ± 5.2 | 35.5 (31.6 – 38.5) | 43.9 | 25.6 | 39.2 ± 6.3 | 38.4 (34.3 – 43.1) | 57.4 | **0.182; 0.003** |
| Whole blood Mn (μg/L) | 4.9 | 8.7 ± 2.8 | 8.4 (6.8 – 9.7) | 16.5 | 6.2 | 10.6 ± 3.3 | 9.9 (8.0 – 12.7) | 19.1 | **0.101; 0.028** |
| Whole blood Na (mg/L)<sup>+</sup> | 998 | 2,042 ± 373 | 2,010 (1800 – 2283) | 2,863 | 1510 | 2,257 ± 384 | 2,286 (1984 – 2569) | 2,842 | **0.108; 0.023** |
| Whole blood Pb (μg/L) | 0.5 | 9.5 ± 8.8 | 8.7 (4.3 – 13.6) | 47.9 | 5.0 | 24.5 ± 19.7 | 20.3 (12.5 – 28.9) | 113 | **0.378; <0.001** |
| Whole blood Se (μg/L)<sup>+</sup> | 66 | 138 ± 50 | 122 (93 – 172) | 227 | 82 | 233 ± 103 | 203 (155 – 296) | 527 | **0.391; <0.001** |
| Whole blood Sr (μg/L)<sup>+</sup> | 8.1 | 14.0 ± 5.2 | 11.8 (10.1 – 16.8) | 30.5 | 5.3 | 13.4 ± 6.4 | 12.8 (9.6 – 14.8) | 45.7 | 0.002; 0.743 |
| Whole blood Zn (μg/L)<sup>+</sup> | 2.8 | 5.3 ± 1.5 | 5.6 (4.1 – 6.5) | 8.1 | 2.9 | 6.6 ± 1.3 | 7.0 (6.0 – 7.4) | 9.0 | **0.180; 0.003** |
| **<u>Welding-related exposure</u>** |  |  |  |  |  |  |  |  |  |
| E90 (mg-days/m<sup>3</sup>) | 0 | 0 ± 0 | 0 (0 – 0) | 0 | 0 | 2.3 ± 2.0 | (0.6 – 2.2) | 7.0 | ---- |
| HrsW (hours) | 0 | 0 ± 0 | 0 (0 – 0) | 0 | 0 | 266 ± 203 | (85 – 360) | 720 | ---- |
| YrsW (years) | 0 | 0 ± 0 | 0 (0 – 0) | 0 | 3 | 25 ± 10 | (18 – 33) | 40 | ---- |
| ELT (mg-years/m<sup>3</sup>) | 0 | 0 ± 0 | 0 (0 – 0) | 0 | 0.17 | 1.14 ± 0.76 | (0.31 – 1.91) | 2.49 | ---- |

Data represent the minimum (min), mean ± standard deviation (SD), median, first and third quartiles (Q1 and Q3, respectively), maximum (max) values, numbers of subjects (n), and percentages (%). Demographic and clinical data were compared using one-way analysis of variance (ANOVA), chi-square, or Mann-Whitney U tests. Whole-blood metal concentrations were compared using analysis of covariance (ANCOVA), controlling for age. p-values and effect sizes ( $\eta_p^2$ ) are reported. Abbreviations: UPDRS-III, Unified Parkinson's Disease Rating Scale-part III (motor scores); E90 = cumulative 90-day exposure to welding fumes; HrsW = hours welding; YrsW = years welding; ELT = lifetime cumulative exposure to welding fumes; Cu, copper; Fe, iron; K, potassium; Mg, magnesium; Mn, manganese; Na, sodium; Pb, lead; Zn, zinc; Se, selenium; Sr, strontium. \*Restricted to current or former smoking or alcohol consumption; <sup>+</sup>Available data for exposed participants = 38.

The proportion of ever-smokers differed significantly, with exposed participants exhibiting a higher frequency of smoking behavior (*p*-value = 0.006). No difference, however, was found in mean smoking pack-years between exposed and non-exposed participants *[U = 64.00, Z = –0.11, p = 0.911]*. Regarding alcohol consumption, the frequency of those consuming any alcohol vs. none did not differ significantly (*p*-value = 0.624). Among those who consumed alcohol, average alcohol intake (drinks per week) did not differ across exposure groups *[U = 69.00, Z = −0.59, p = 0.557]*. On average, exposed participants had welded for approximately 25 years, and their average hours worked in the past 90 days were approximately 266 (Table 1).

### Whole-blood metal concentrations in exposed and non-exposed

The ANCOVA revealed that compared to non-exposed, exposed participants had significantly higher concentrations of several metals in whole blood. These were: Cu *[F(1,66) = 26.00, p-value < 0.001]*; Fe *[F(1,66) = 9.55, p-value = 0.003]*, K *[F(1,66) = 29.33, p-value < 0.001]*, Mg *[F(1,65) = 10.21, p-value = 0.003]*, Mn *[F(1,66) = 5.14, p-value = 0.028]*, Na *[F(1,67) = 5.55, p-value = 0.023]*, Pb *[F(1,67) = 27.98, p-value < 0.001]*, Zn *[F(1,65) = 10.06, p-value = 0.003]*, and Se *[F(1,67) = 29.49, p-value <0.001]* (Table 1).

### Exposure-based differential expression analysis of miRNAs

To evaluate the effect of metal mixture exposure on circulating miRNA expression, we conducted next-generation RNA sequencing on serum-derived EVs from non-exposed and exposed participants. A total of 1,070 known human miRNAs and 13 PIWI-interacting RNAs (piRNAs) were detected across all samples. After filtering out miRNAs and piRNAs with <10 read counts in <3 samples, the remaining data were log-transformed for downstream analysis.

Differential expression analysis proceeded in two steps: first, miRNAs with false discovery rate (FDR)–adjusted p-values < 0.05 were identified as differentially expressed; second, to facilitate biological interpretation, a fold-change reference threshold (absolute log2FC ≥ 0.8) was applied to prioritize miRNAs with larger effect sizes. Using these criteria, we identified 50 miRNAs and 1 piRNA as significantly differentially expressed, with 37 being greater-expressed and 14 lesser-expressed in exposed compared with non-exposed participants (Supplementary Table 1). Further analyses were performed on this final set of 50 miRNAs and 1 piRNA.

Hierarchical clustering heatmap and volcano plots analyses were performed to visualize overall miRNA expression differences between non-exposed and exposed participants for these 50 miRNAs and 1 piRNA with an adjusted *p*-value < 0.05 (Supplementary Figures S1 and S2). The heatmap shows that non-exposed and exposed participants cluster into separate groups, except for 9 non-exposed and 3 exposed participants. A similar pattern emerged from the PCoA analysis (Supplementary Figure S2 Panel A) that also found a significant difference in miRNA expression between non-exposed and exposed participants. In this latter analysis, the PERMANOVA test was significant (adjusted *p*-value = 0.007), whereas the permutation test was non-significant (adjusted *p*-value = 0.284). This indicates a significant difference in the distribution between non-exposed and exposed participants, and also that the dispersion did not cause the difference. Lastly, the LEfSe plot, a method used for metagenomic biomarker discovery, detected the miRNAs with the greatest differential expression in the two exposure groups (Supplementary Figure S2 Panel C).

### Exposure–miRNA associations

Of the 50 miRNAs and 1 piRNA that were differentially expressed between non-exposed and exposed participants, *Pearson* partial correlation analyses adjusted for age and subsequently for both age and exposure group identified 23 miRNAs significantly associated with at least one whole-blood metal concentration across the cohort (p<0.05). As part of our exploratory screening approach, we subsequently prioritized miRNAs that: 1) were significantly associated with more than one of the 10 quantified metals; 2) showed at least one correlation of moderate magnitude (|r| ≥ 0.30); and 3) exhibited consistent directionality across multiple neurotoxic metals.

This approach resulted in a subset of 12 miRNAs (miR-16-5p, miR-22-3p, miR-3150a-3p, miR-378f, miR-4417, miR-4646-3p, miR-486-5p, miR-6746-5p, miR-6775-3p, miR-6857-3p, miR-8056, and miR-93-5p). The detectability rates, however, varied across these miRNAs limiting their application for continuous exposure–response modeling.

Our multi-criteria approach incorporated: (i) differential expression; (ii) multi-metal associations; (iii) magnitude and directional consistency of correlations; and (iv) appropriate distribution for continuous exposure–response modeling (expression detectable in nearly all participants). After using converging epidemiological, experimental and neuropathological evidence that links miRNAs to metal exposure and neurodegenerative mechanisms, three miRNAs remained (**miR-16-5p**, **miR-93-5p**, and **miR-486-5p**). These miRNAs showed consistent cross-metal association patterns in our dataset and have been independently reported as implicated in metal exposure and neurodegenerative conditions including ADRD and PD (Supplementary Table S2). They were therefore prioritized for subsequent exposure–response and metal mixture modeling analyses.

*Pearson* partial correlation analysis revealed negative associations between miR-16-5p, miR-93-5p, and miR-486-5p expression and whole-blood concentrations of metals particularly Pb, Se, and Cu (Table 2). After applying FDR correction across the three prioritized miRNAs and 10 metals, only the association between Pb and miR-16-5p remained statistically significant (q = 0.030). The data distributions (unadjusted and log-transformed) and corresponding linear trends are shown in Supplementary Figure S3.

**Table 2.** Pearson partial correlation between miRNAs, whole-blood metals and welding-related exposure measures in the total cohort (n=66).

| miRNA | Correlation | Values | Whole-blood metal |  |  |  | Welding-related exposure measures |  |  |  |
| --- | --- | --- | --- | --- | --- | --- | --- | --- | --- | --- |
|  |  |  | Cu | Pb | Se | K | YrsW | ELT | HrsW | E90 |
| miR-16-5p | Adj. age | R | -0.46 | -0.53 | -0.49 | -0.39 | -0.41 | -0.32 | -0.25 | -0.25 |
|  |  | P | 0.011 | 0.004 | 0.004 | 0.043 | 0.002 | 0.011 | 0.171 | 0.227 |
|  | Adj. age, group | R | -0.28 | -0.37 | -0.32 | -0.17 | -0.20 | -0.10 | 0.17 | 0.16 |
|  |  | P | 0.024 | 0.003 | 0.011 | 0.180 | 0.125 | 0.417 | 0.177 | 0.193 |
| miR-93-5p | Adj. age | R | -0.38 | -0.51 | -0.44 | -0.45 | -0.49 | -0.43 | -0.35 | -0.33 |
|  |  | P | 0.056 | 0.004 | 0.011 | 0.006 | 0.002 | 0.003 | 0.038 | 0.049 |
|  | Adj. age, group | R | -0.15 | -0.33 | -0.23 | -0.24 | -0.11 | -0.15 | 0.07 | 0.04 |
|  |  | P | 0.247 | 0.007 | 0.070 | 0.053 | 0.389 | 0.235 | 0.575 | 0.759 |
| miR-486-5p | Adj. age | R | -0.49 | -0.46 | -0.44 | -0.36 | -0.43 | -0.31 | -0.24 | -0.22 |
|  |  | P | 0.005 | 0.015 | 0.012 | 0.062 | 0.002 | 0.015 | 0.171 | 0.228 |
|  | Adj. age, group | R | -0.31 | -0.29 | -0.25 | -0.14 | -0.17 | -0.05 | 0.19 | 0.20 |
|  |  | P | 0.012 | 0.026 | 0.046 | 0.281 | 0.190 | 0.682 | 0.136 | 0.187 |

Abbreviations: miRNA, microRNA; R, correlation coefficient (adjusted for age or age + exposure group); P, p-value (adjusted for age or age + exposure group); Cu, copper; Pb, lead; Se, selenium; K, potassium; YrsW, years welding; ELT, lifetime cumulative welding exposure; E90, cumulative 90-day welding exposure; HrsW, hours welding.

*Pearson* partial correlation analyses were conducted between the three prioritized miRNAs (miR-16-5p, miR-93-5p, and miR-486-5p) and welding-related exposure measures (YrsW, ELT, HrsW and E90). In models adjusted for age alone, long-term exposure metrics were negatively associated with miRNA expression (Table 2). After additional adjustment for exposure groups, however, these associations were no longer statistically significant, suggesting that the initial findings were largely driven by exposure groups contrast (exposed vs. non-exposed). The data distributions (unadjusted and log-transformed) and corresponding linear trends are shown in Supplementary Figure S4.

### Metal mixture modeling: BKMR analysis

BKMR was applied to evaluate the relative importance of individual metals within the mixture on the expression of the three prioritized miRNAs (miR-16-5p, miR-93-5p, and miR-486-5p), initially adjusting for age. All models additionally adjusted for exposure groups are presented in Supplementary Figures S5–S10.

Posterior inclusion probabilities (PIPs) indicated the contribution of each metal to the model (Table 3).Whole-blood Pb consistently exhibited the highest PIPs across models for miR-16-5p, miR-93-5p, and miR-486-5p suggesting that Pb was the predominant component of the metal mixture associated with miRNA expression. After additional adjustment for both age and exposure group Pb remained to exhibit the highest PIPs, particularly for miR-16-5p, miR-93-5p, although the magnitude of the PIPs was reduced.

**Table 3.** Posterior inclusion probabilities (PIPs) for individual metals (natural log-transformed) in BKMR model predicting miRNA expression. Models were adjusted for age (left) and for both age and exposure group (right). PIPs reflect the relative importance of each metal in the model.

| *Whole-blood metals* | *Posterior Inclusion Probability (PIP)*<br>*Adj. age* |  |  | *Posterior Inclusion Probability (PIP)*<br>*Adj. age and exposure group* |  |  |
| --- | --- | --- | --- | --- | --- | --- |
|  | **miR-16-5p** | **miR-93-5p** | **miR-486-5p** | **miR-16-5p** | **miR-93-5p** | **miR-486-5p** |
| ***Cu*** | 0.49 | 0.52 | 0.70 | 0.50 | 0.49 | 0.59 |
| ***Fe*** | 0.49 | 0.50 | 0.61 | 0.53 | 0.54 | 0.57 |
| ***K*** | 0.44 | 0.57 | 0.57 | 0.51 | 0.56 | 0.51 |
| ***Mg*** | 0.47 | 0.56 | 0.55 | 0.43 | 0.55 | 0.54 |
| ***Mn*** | 0.39 | 0.49 | 0.60 | 0.42 | 0.53 | 0.53 |
| ***Pb*** | **0.84** | **0.98** | **0.86** | **0.68** | **0.73** | **0.56** |
| ***Zn*** | 0.40 | 0.60 | 0.55 | 0.41 | 0.53 | 0.51 |
| ***Na*** | 0.42 | 0.52 | 0.56 | 0.47 | 0.50 | 0.50 |
| ***Se*** | 0.63 | 0.58 | 0.59 | 0.51 | 0.52 | 0.50 |
| ***Sr*** | 0.39 | 0.52 | 0.57 | 0.40 | 0.43 | 0.55 |

*Abbreviations: Copper (Cu), Iron (Fe), Potassium (K), Magnesium (Mg), Manganese (Mn), Lead (Pb), Zinc (Zn), Sodium (Na), Selenium (Se), and Strontium (Sr).*

The overall mixture effect suggested an inverse association between the metal mixture and miRNA expression (Supplementary Figure 5). Higher quantiles of the metal mixture were progressively associated with decreased expression of miR-16-5p, miR-93-5p, and miR-486-5p, particularly above the 60^th^ percentile. After additional adjustment for exposure group, the direction of the associations remained consistent, however, the 95% credible intervals crossed zero, suggesting that part of the overall mixture effect reflects the contrast between groups.

Single exposure–response associations indicated that higher whole-blood Pb levels were consistently associated with decreased expression of miR-16-5p, miR-93-5p (Figure 2A and 2B), and miR-486-5p (Figure 2C), while holding all other metals at their median (50^th^ percentile). The inverse association persisted across the upper Pb range. In contrast, the exposure–response curves for the remaining metals were largely flat across the exposure range, and the 95% credible intervals crossed zero, suggesting minimal contributions within the mixture model. After additional adjustment for exposure group, the Pb–miRNA associations remained directionally consistent.

**Figure 2.**
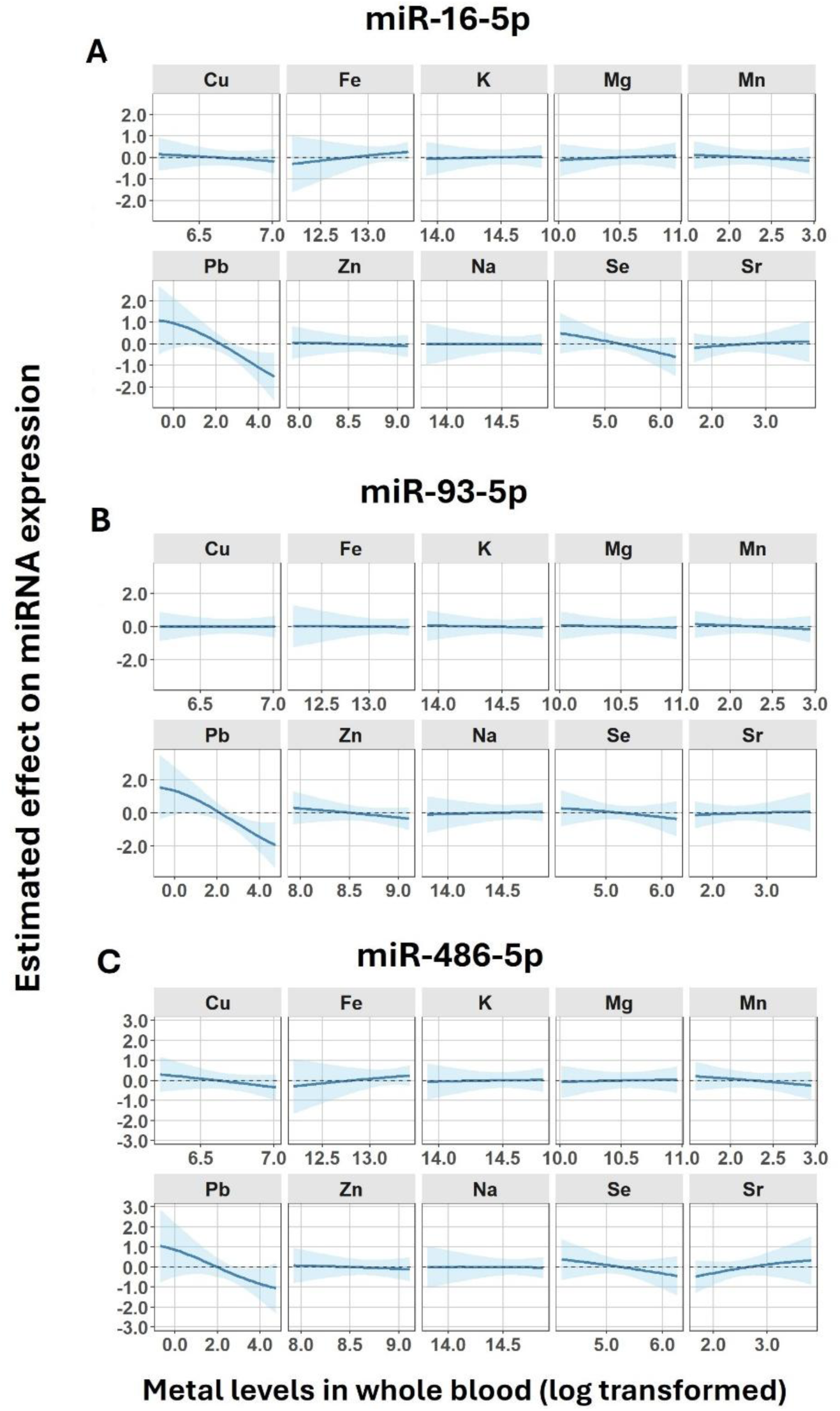
Single-metal exposure–response between individual log-transformed metal levels in whole blood and log-transformed expression levels of: A) miR-16-5p, B) miR-93-5p, and C) miR-486-5p. Blue lines indicate mean estimated effects, and shaded areas represent the 95% credible intervals, holding all other metals at their 50th percentile. Metals included: copper (Cu), iron (Fe), lead (Pb), magnesium (Mg), manganese (Mn), potassium (K), selenium (Se), sodium (Na), strontium (Sr), and zinc (Zn).

Bivariate exposure–response further supported the negative association between whole-blood Pb levels and miRNA expression after adjusting for age for miR-16-5p (Figure 3), miR-93-5p (Figure 4), and miR-486-5p (Figure 5), across percentiles of co-exposure metals (Pb as expo1). Notably, the negative association of Pb was amplified in the presence of higher Se levels (Figure 3-5). Stratified analyses (data not shown), however, indicate that this Se × Pb interaction was primarily driven by non-exposed, with no effect observed among exposed participants. When Pb was held constant (Pb as expos2), estimated effects of remaining metals varied across Pb quantiles. After additional adjustment for exposure group, the Pb–miRNAs association remained consistent across different exposure percentiles.

**Figure 3.**
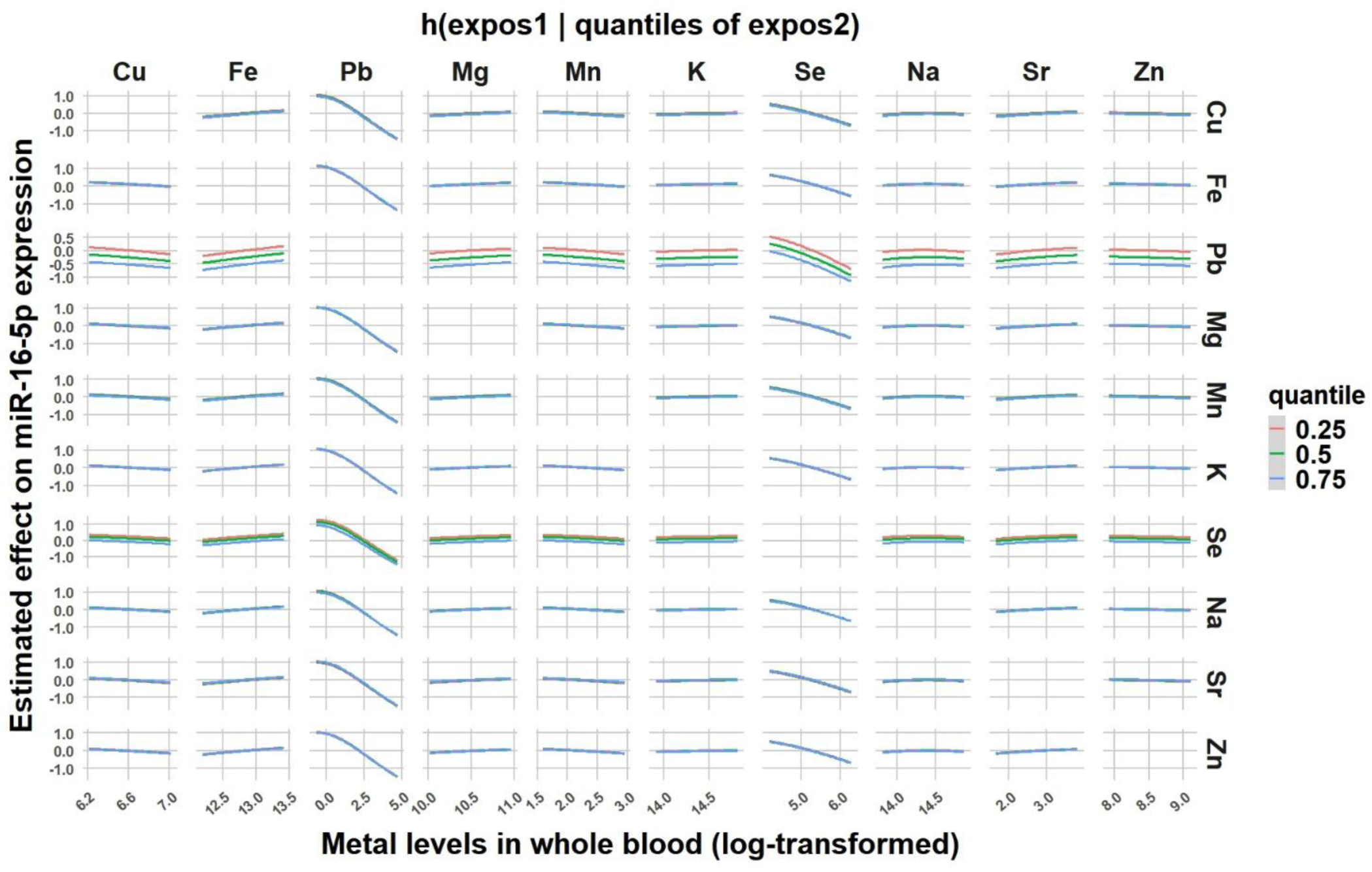
Bivariate exposure–response between individual log-transformed metal levels in whole blood (x-axis; expo 1) and log-transformed expression levels of: miR-16-5p conditional on another metal (expo 2) held at the 25th (red), 50th (green), or 75th (blue) percentiles. Metals included: copper (Cu), iron (Fe), lead (Pb), magnesium (Mg), manganese (Mn), potassium (K), selenium (Se), sodium (Na), strontium (Sr), and zinc (Zn).

**Figure 4.**
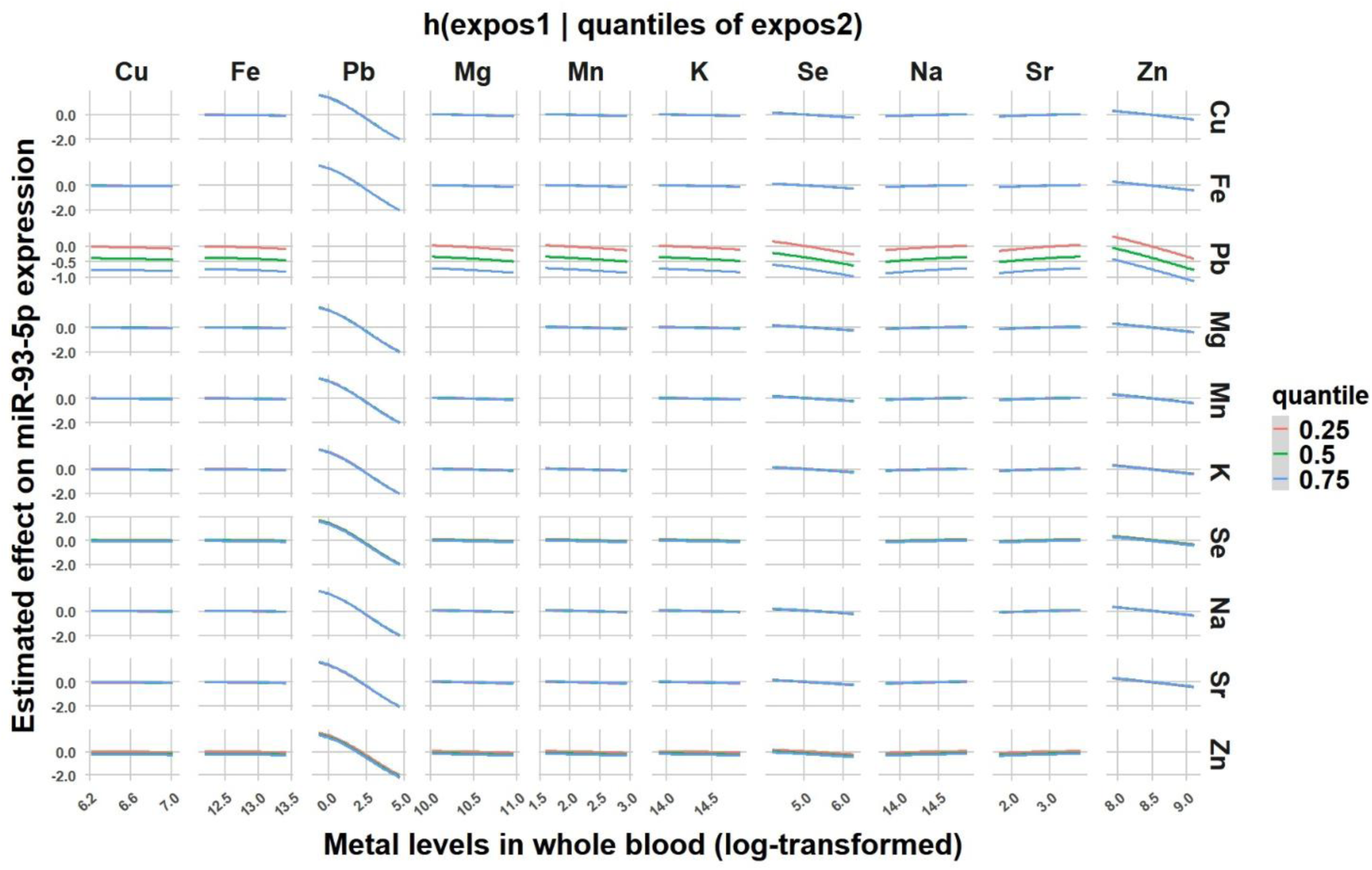
Bivariate exposure–response between individual log-transformed metal levels in whole blood (x-axis; expo 1) and log-transformed expression levels of: miR-93-5p conditional on another metal (expo 2) held at the 25th (red), 50th (green), or 75th (blue) percentiles. Metals included: copper (Cu), iron (Fe), lead (Pb), magnesium (Mg), manganese (Mn), potassium (K), selenium (Se), sodium (Na), strontium (Sr), and zinc (Zn).

**Figure 5.**
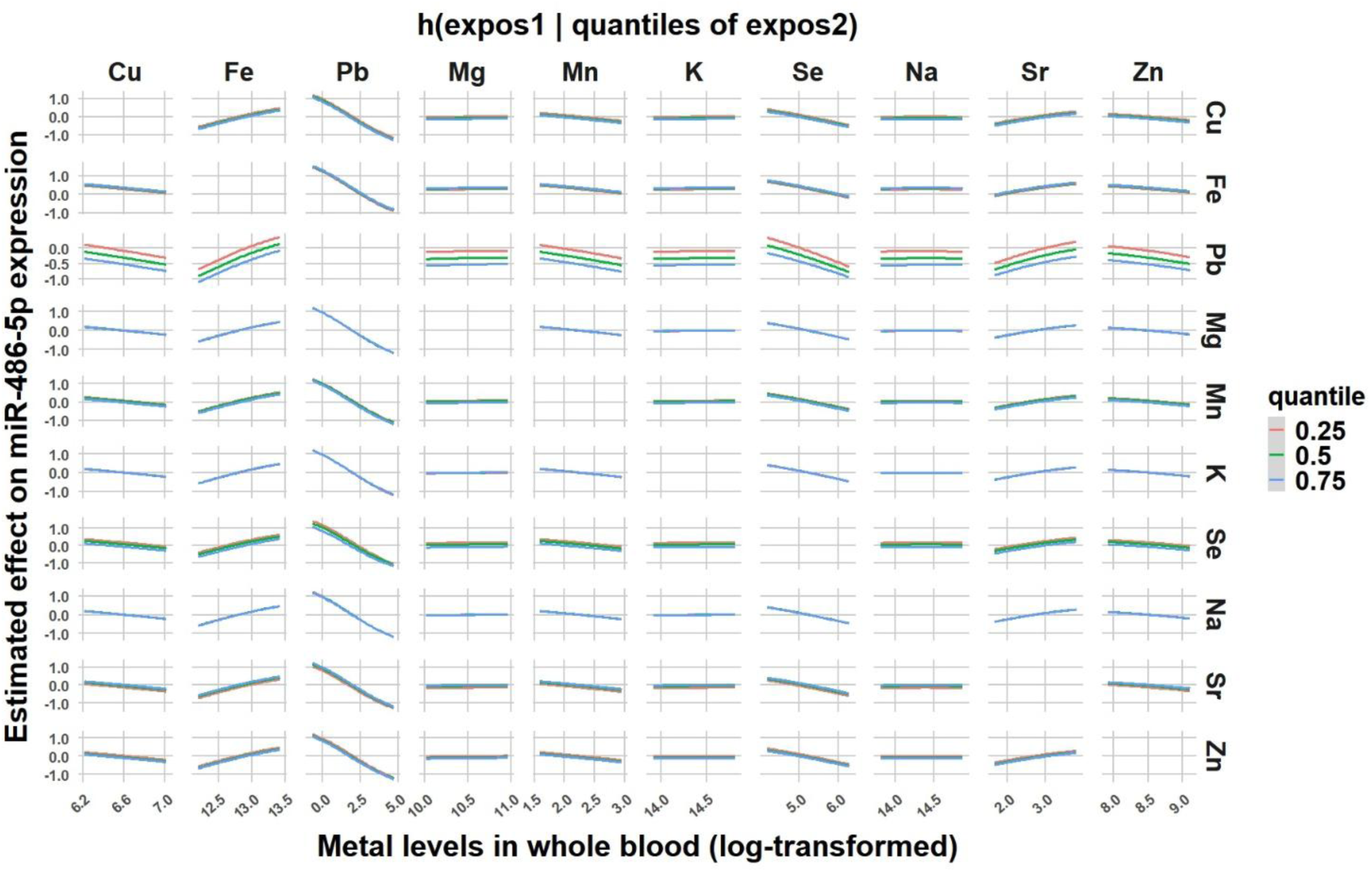
Bivariate exposure–response between individual log-transformed metal levels in whole blood (x-axis; expo 1) and log-transformed expression levels of: miR-486-5p conditional on another metal (expo 2) held at the 25th (red), 50th (green), or 75th (blue) percentiles. Metals included: copper (Cu), iron (Fe), lead (Pb), magnesium (Mg), manganese (Mn), potassium (K), selenium (Se), sodium (Na), strontium (Sr), and zinc (Zn).

Lastly, single-variable risk analysis indicated that increasing Pb levels from the 25^th^ to 75^th^ percentile, whereas holding other metals fixed at specific quantiles was associated with significant reduced expression of miR-16-5p (Δy [P25-P75] = −0.54; 95% CrI: [−0.94,-0.15]) and miR-93-5p (Δy [P25-P75] = −0.74; 95% CrI: [−1.24, −0.25]), after adjusting for age (Figure 6). In contrast, no statistical significance was found for miR-486-5p (Δy [P25–P75] ≈ −0.22; 95% CrI: [−0.591, 0.142]). After additional adjustment for exposure group, the association remained significant for miR-16-5p (Δy [P25-P75] ≈ −0.38; 95% CrI: [−0.75,-0.01]), whereas the estimate for miR-93-5p was attenuated (Δy [P25–P75] ≈ −0.41; 95% CrI: [−0.84, 0.02]).

**Figure 6.**
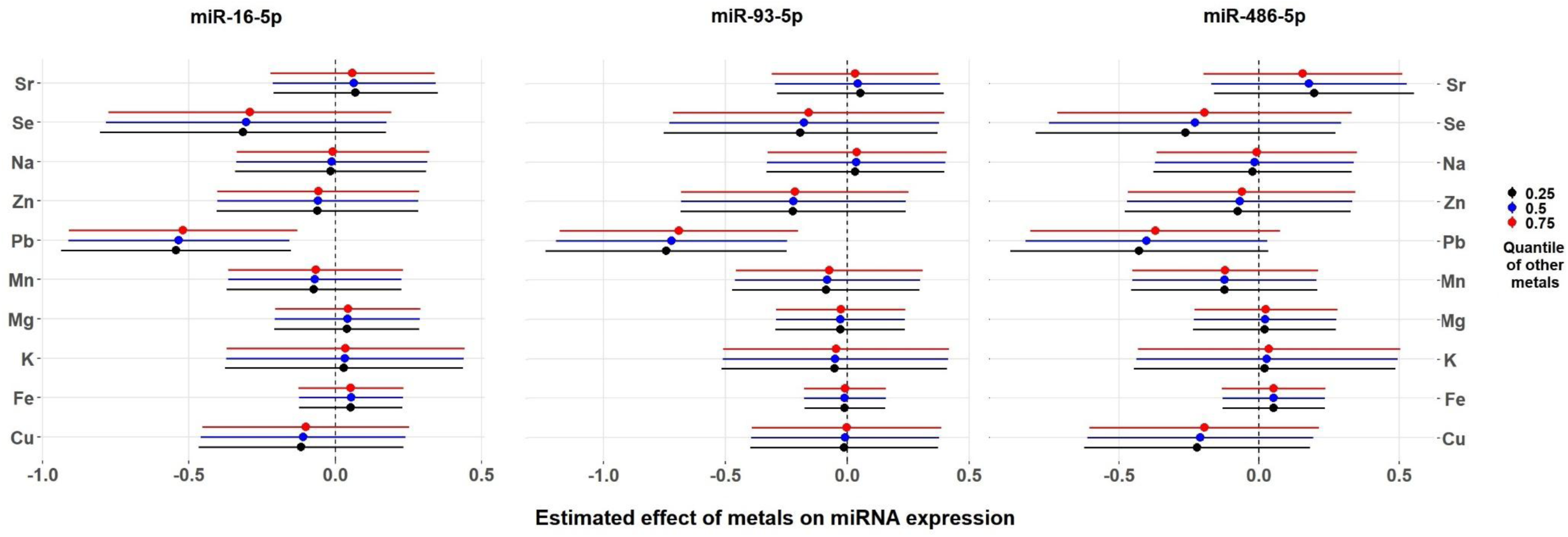
Single-variable risk for each metal on expression levels of: miR-16-5p (left), miR-93-5p (center), and miR-486-5p (right), conditional on other metals held at the 25^th^, 50^th^ and75th percentile (black, blue, and red, respectively).

### Exploratory mediation linking Pb-associated miRNAs and brain MRI R2* measure

Because miR-16-5p and miR-93-5p were consistently associated with whole-blood Pb levels, we next explored whether these miRNAs were also related to R2* MRI measures. Given prior evidence linking Pb exposure to R2* in the red nucleus (RN) in this cohort^4^, and significant correlations observed only for the RN (Supplementary Table S3), we examined whether miR-16-5p and miR-93-5p independently mediate the association between whole-blood Pb levels and R2* RN (Figure 7).

**Figure 7.**
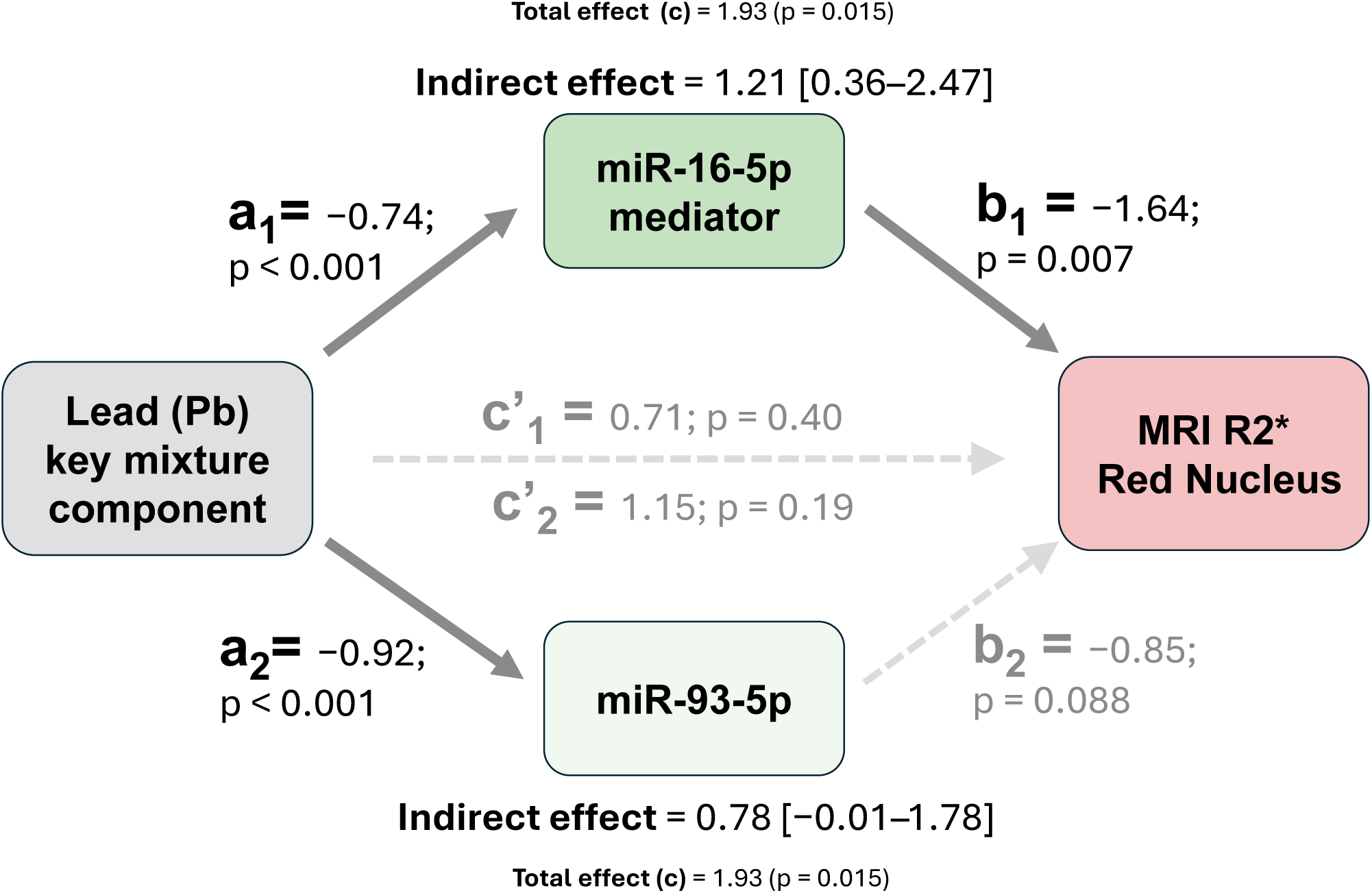
Mediation analysis linking whole-blood lead (Pb) exposure, circulating neuron-derived miRNAs, and iron-sensitive MRI signal (R2*) in the red nucleus (RN). Path coefficients are shown for models adjusted for age. Higher Pb levels were associated with reduced circulating miRNA expression for miR-16-5p (path a1) and miR-93-5p (path a2). Lower miR-16-5p expression was significantly associated with higher R2* values in the red nucleus (path b1), whereas the association for miR-93-5p showed a similar direction but did not reach statistical significance (path b2). Mediation analysis indicated a significant indirect effect of Pb on R2* RN through miR-16-5p (β = 1.21, 95% CI [0.36–2.47]), while the indirect effect through miR-93-5p was not significant (β = 0.78, 95% CI [−0.01–1.78]). The dashed horizontal path represents the direct association between Pb exposure and R2* RN after inclusion of the mediator in the model (c′1 for miR-16-5p; c’2 for miR-93-5p). The total effect of Pb on R2* RN was significant (β = 1.93, p = 0.015).

Higher whole-blood Pb levels were associated with lower miR-16-5p expression (β = −0.74, p < 0.001), and lower miR-16-5p expression was associated with higher R2* RN values (β = −1.64, p = 0.007). Mediation analysis indicated a significant indirect effect of Pb on R2* RN through miR-16-5p (β = 1.21, 95% CI [0.36–2.47]) (Figure 7), which remained significant after additional adjustment for exposure group (Supplementary table S4). After inclusion of miR-16-5p in the model, the direct Pb–R2* RN association was no longer significant, consistent with an indirect-only mediation pattern (Figure 7), suggesting that the association between Pb exposure and R2* RN may occur through miR-16-5p. miR-93-5p showed a similar direction of association but the indirect effect did not reach statistical significance (Figure 7). Full mediation results adjusting for age and subsequently for age and group are presented in Supplementary table S4.

## Discussion

In this study, we investigated whether circulating neuron-derived miRNAs could reflect molecular perturbation associated with metal mixtures exposure. While differential expression analyses identified 50 dysregulated miRNAs between exposure groups, mixture modeling indicated that Pb consistently emerged as the metal most strongly associated with reduced expression of specific miRNAs, particularly miR-16-5p and miR-93-5p. These findings suggest that within complex metal mixtures, molecular signals may be related to specific metals known to be toxic, and support circulating neuron-derived miRNAs as early indicators of exposure-related neurobiological vulnerability.

### Biological meaning of neuron-derived EVs

The miRNA profiles analyzed in this study derived from neuron-enriched EVs isolated from peripheral blood, providing a minimally invasive window into brain-related molecular processes. Given their role in post-transcriptional gene regulation, circulating miRNAs may reflect molecular regulatory networks perturbation influenced by environmental exposures,^21^ and serve as accessible indicators of CNS-related toxicity before clinical symptoms emerge.^21,22^

In our cohort, exposed participants exhibited a particular panel of differentially expressed miRNAs. Reduced expression of miR-16-5p, miR-93-5p, and miR-486-5p, implicated in oxidative stress regulation,^37,38^ inflammatory signaling,^39,40^ and cellular survival, may reflect subclinical neurobiological perturbation associated with exposure to metal mixtures. These miRNAs also play complementary neuroprotective roles by modulating microglia-mediated neuroinflammation,^41–44^ neurogenesis and neuronal growth.^45^ Notably, reduced expression of these miRNAs has been observed in several NDDs, including PD,^28^ AD,^46–48^ young-onset Alzheimer’s disease (YOAD),^13^ severe dementia [sporadic Creutzfeldt-Jakob disease (CJD)],^48,49^ and AD progression.^15^ Although participants in our study were asymptomatic, these convergent molecular patterns suggest that metal mixtures exposure may influence regulatory molecular pathways relevant to neurobiological vulnerability.

In human studies, causality is difficult to prove, but our findings are consistent with large-scale exposome studies indicating that circulating molecular signatures in blood, including miRNAs, may capture biological processes related to environmental toxicants.^21,22^ To our knowledge, this is the first study to profile circulating miRNAs from neuron-derived extracellular vesicles in relation to metal mixtures, highlighting their potential to capture early molecular signatures of neurobiological stress related to metals co-exposure.

### Lead as a key contributor to miRNA perturbation within the metal mixture

Our analytical approach allowed us to explore the associations between mixed-metal exposure and miRNA expression accounting for nonlinear and potential interaction effects among metals, providing a more realistic representation of real-word exposure. Adjustment for exposure group attenuated some associations, suggesting that part of the observed findings reflect the exposure contrast introduced by the study design. Importantly, the direction of the associations persisted after exposure group adjustment, including significant association between Pb and miR-16-5p and a similar trend for miR-93-5p. These observations as consistent with previous studies reporting reduced miR-93-5p expression associated with elevated patellar Pb levels in a population-based sample,^50^ and miR-16-5p reduced in metal-exposed workers, particularly in the presence of complex mixtures.^12^

Our findings suggest that the molecular patterns associated with metal co-exposure may reflect distinct contribution of individual metals within the mixture, with Pb emerging as the most relevant component in this cohort. This observation aligns with metal mixtures exposure scenarios, where multiple metals coexist but the biological effect may be driven by specific components within the mixture.^12,51^ Despite the modest sample size, we provide converging evidence from DE analysis, exposure–miRNA associations, and mixture modeling suggesting that Pb may represent a key component of the metal mixture associated with neuron-derived circulating miRNAs.

Pb is a well-established neurotoxicant, even at relatively low exposure levels,^4,52^ and has been associated with decreased global DNA methylation in occupationally exposed workers,^53^ with neurotoxic effects,^4,52,54,55^ and with increased risk of neurodegenerative conditions.^14,56–58^ Previous studies have also reported dysregulation of inflammation-related miRNAs, such as miR-155, in individuals with high Pb exposure^59^ and in occupationally exposure setting.^60^ Collectively, this evidence support the biological relevance of Pb exposure on molecular perturbation^6,52,61^ and its potential involvement on stress- and inflammation-related regulatory pathways.^12,60^ Our findings extend this evidence by demonstrating similar associations in neuron-derived circulating miRNAs, suggesting that these peripheral molecular signature may reflect early regulatory processes in the brain associated with Pb exposure.

### Biological pathways underlying Pb exposure to miRNA perturbation

Pb exposure is well known to induce oxidative stress^6,61^ and disrupt metal homeostasis in neural tissue.^6^ Such disturbances can alter brain iron regulation and promote local accumulation in vulnerable regions of the brain.^4^ These processes may also influence the regulation of stress-responsive miRNAs through mechanisms including oxidative stress signaling, activation of pro-inflammatory cytokines (TNF-α, IL-1β, and IL-6),^5^ and epigenetic alterations affecting miRNA transcription.^53,62–64^

In this context, the reduced expression of miR-16-5p and miR-93-5p observed in our cohort may reflect biological mechanisms through which Pb alters miRNA regulation and cellular homeostasis. miR-16-5p and miR-93-5p regulate signaling pathways involved in inflammation, cellular stress responses, and survival, including PI3K–AKT/mTOR signaling, p53-mediated apoptosis, autophagy, and MAPK activation.^65–67^ Together with miR-486-5p, these miRNAs converge on regulatory networks implicated in mitochondrial homeostasis, oxidative stress modulation, and neuronal survival, processes frequently disrupted by Pb exposure.^5,6,14^ These molecular mechanisms provide a potential framework connecting Pb exposure to perturbations in neuron-derived circulating miRNAs and MRI signals.

### Translational relevance of circulating miRNAs for brain iron-related MRI signals

Our analyses were initially designed to identify circulating neuron-derived miRNAs associated with metal exposure, among which Pb emerged as the metal most consistently associated with these miRNAs. Because whole-blood Pb levels were consistently associated with miR-16-5p and miR-93-5p, we next explored whether these miRNAs were also related to R2* MRI measures. Given our previous study linking Pb exposure to R2* in the red nucleus (RN) in this cohort^4^, and significant correlations observed only for the RN (Supplementary Table S3), we examined whether miR-16-5p and miR-93-5p independently mediate the Pb-R2* RN association between whole-blood Pb levels and (Figure 6). Interestingly, our mediation analyses indicated a potential mediating role of miR-16-5p linking Pb exposure to R2* in the red nucleus.

Members of the miR-16 family regulate Divalent Metal Transporter-1 (DMT1) and other pathways involved in inflammatory signaling, and cellular survival, supporting the possibility that reduced miR-16-5p expression may relate to altered iron handling.^68^ Because R2* MRI measures are sensitive to magnetic susceptibility effects related to iron,^36^ alterations in R2* signal may reflect cellular responses associated with disrupted iron handling and oxidative stress.^69^ Increased R2* values in this region have been associated with higher glial cell density and alterations in iron regulation, suggesting that susceptibility MRI signals may capture cellular responses related to glial activation, neuroinflammatory processes, and oxidative stress.^69^The red nucleus is an iron-rich brain structure with high oxidative metabolic activity that make it particularly vulnerable to such perturbations.^69^ Consistent with this biological vulnerability, increased R2* values in the red nucleus among individuals with higher Pb exposure,^1^ and similar alterations have been reported in neurodegenerative conditions such as Parkinson’s disease.^70^ Together, these observations suggest that reduced miR-16-5p expression may capture molecular pathways related to iron transport and homeostasis, providing potential link between Pb exposure and iron-sensitive MRI signals in the brain.

miR-16-5p and miR-93-5p belong to distinct microRNA families but participate in overlapping regulatory networks. The consistent directionality observed for both miRNAs in our analyses suggests that these molecules may capture related biological processes triggered by metal exposure. miR-93-5p showed a similar directionality, but with weaker statistical support. These findings indicate that miR-16-5p may represent a more robust molecular mediator linking Pb exposure to iron-sensitive MRI signals, while miR-93-5p may reflect related biological processes within overlapping regulatory networks involved in cellular stress responses. Further studies integrating molecular, imaging, and mechanistic approaches are needed to clarify the biological pathways linking metal exposure, circulating miRNAs, and iron-related brain accumulation.

### Limitations, implications, and future directions

This comprehensive analysis in asymptomatic individuals yielded important insights, but our study has several limitations. First, the sample size was relatively small, and only male participants were included. Second, the variability in exposure levels and the composition of metal mixtures may influence miRNA expression. Additionally, the cross-sectional design makes it impossible to claim direct causal relationships. Future longitudinal studies that also assesses sex differences will be important to address mechanisms and overall impact of chronic metal exposure on miRNA dynamics.

### Conclusion

In summary, our findings indicate that individuals with higher metal exposure exhibit a distinct neuron-derived circulating miRNA profile associated with metal mixtures. Reduced expression of miR-16-5p, miR-93-5p, and miR-486-5p appear to reflect early subclinical perturbations in molecular pathways related to neuroprotection, oxidative stress, and inflammation. Within the mixture, Pb emerged as the metal most consistently associated with these miRNAs, suggesting that they may serve as molecular signatures linking Pb exposure to iron-sensitive brain MRI signals. Collectively, these findings support circulating neuron-derived miRNAs as minimally invasive markers of exposure-related neurobiological processes and highlight the translational potential of blood-based biomarkers for monitoring early environmental impacts on brain health.

## Data Availability

Data not included in manuscript or supplemental information is available to qualified investigators upon request.

## Acknowledgements

We would like to thank all the volunteers who participated in this study. We are also indebted to many individuals who helped make this work possible, including Melissa Santos and Susan Kocher for subject coordination, recruitment, blood sample handling, and data entry; Michael R. Flynn, Rebecca Fry, and Lisa Smeester (Department of Environmental Sciences and Engineering, University of North Carolina at Chapel Hill) for their contributions to the study design and generation of the exposure data; Pam Susi and Pete Stafford of CPWR; Mark Garrett, John Clark, and Joe Jacoby of the International Brotherhood of Boilermakers; Fred Cosenza and all members of the Safety Committee for the Philadelphia Building and Construction Trades Council; Ed McGehean of the Steamfitters Local Union 420; Jim Stewart of the Operating Engineers; Sean Gerie of the Brotherhood of Maintenance of Way Employees Division, Teamsters Rail Conference; and Terry Peck of Local 520 Plumbers, Pipefitters, and HVAC.

Mechelle M. Lewis participated in this research when employed at Penn State University. The opinions expressed in this article are the author’s own and do not reflect the view of the U.S. National Institutes of Health.

## Funding

This work was supported by National Institutes of Health grant R01 ES019672. This work was also supported by the U.S. Army Medical Research Materiel Command, endorsed by the U.S. Army through the Parkinson’s Research Program (PRP), Investigator-Initiated Research Award (IIRA), Program Announcement Funding Opportunity Announcement Number W81XWH-17-PRR-IIRA, under the award No. W81XWH1810106 to AGK and XH, and University of Virginia Translational Research Development Funds. Other sources include Johnny Isakson Endowment and the Coach Mark Richt Neurological Disease Research Fund to AGK and AK.

## Conflicts of interest

The authors declare no conflicts of interest.

## Author contributions

Conceptualization: JMP-R, AB-C, XH, and AGK;

Analysis and Data curation: JMP-R, AB-C, YIK, ALGR; Methodology, AB-C, JMP-R, JC, VA, YIK;

Project administration, Resources, Funding acquisition: AGK, XH, RBM, MAH, ALGR, AK, VA, PM, MML, HJ, and GZ;

Original draft preparation: JMP-R, AB-C, JDY;

Review and editing, JMP-R, AB-C, YIK, JC, ALGR, MAH, JDY, GZ, HJ, VA, MML, RBM, XH, and AGK.

## Notes

### Competing Interest Statement

The authors have declared no competing interest.

### Funding Statement

This study was by National Institutes of Health grant number R01 ES019672 and by the DOD CMRP.

### Author Declarations

The protocol was reviewed and approved by the Penn State Hershey Institutional Review Board. Written informed consent was obtained from all participants, and the study was conducted consistent with the Declaration of Helsinki.

### Summary of Updates

There was additional analysis and additional data added. The submission was revised accordingly. The title was changed.

