## Supplemental Material for "Neuron-derived circulating miRNAs reveal lead (Pb) as a key component of metal mixtures exposure"

### *S*UPPLEMENTARY *M*ATERIAL *T*ABLE *S1:*

*Differentially expressed small non-coding RNAs (sncRNAs) between non-exposed (n=27) and exposed (n=39) participants.*

| *sncRNA* | *log₂ FC* | *SE* | *Wald statistic* | *Adj. p-values* |
|---|---|---|---|---|
| ***Greater-expression*** | | | | |
| miR-203b-5p | 1.2 | 0.34 | 3.39 | 0.020 |
| miR-205-5p | 1.5 | 0.33 | 4.49 | <0.001 |
| miR-142-5p | 1.6 | 0.44 | 3.67 | 0.012 |
| miR-29c-3p | 1.6 | 0.47 | 3.51 | 0.015 |
| miR-4775 | 1.8 | 0.56 | 3.12 | 0.039 |
| miR-718 | 1.8 | 0.50 | 3.54 | 0.014 |
| miR-4646-3p | 2.0 | 0.60 | 3.38 | 0.020 |
| miR-4632-5p | 2.1 | 0.68 | 3.05 | 0.045 |
| miR-4501 | 2.2 | 0.65 | 3.30 | 0.025 |
| miR-6857-3p | 2.3 | 0.49 | 4.63 | <0.001 |
| miR-4281 | 2.3 | 0.76 | 3.08 | 0.043 |
| miR-6717-5p | 2.5 | 0.72 | 3.50 | 0.015 |
| let-7g-5p | 2.5 | 0.67 | 3.79 | 0.009 |
| miR-1260b | 2.6 | 0.84 | 3.05 | 0.045 |
| miR-4417 | 2.8 | 0.76 | 3.63 | 0.013 |
| miR-3182 | 2.9 | 0.72 | 3.99 | 0.004 |
| miR-4707-5p | 3.0 | 0.58 | 5.25 | <0.001 |
| miR-6808-5p | 3.2 | 0.70 | 4.54 | <0.001 |
| miR-8056 | 3.2 | 0.87 | 3.66 | 0.012 |
| miR-19b-3p | 3.6 | 1.10 | 3.26 | 0.027 |
| miR-5000-5p | 3.6 | 1.03 | 3.51 | 0.015 |
| miR-103b | 3.8 | 1.01 | 3.76 | 0.009 |
| miR-6746-5p | 3.8 | 1.22 | 3.11 | 0.040 |
| miR-653-5p | 3.9 | 1.28 | 3.09 | 0.042 |
| miR-3150a-3p | 4.0 | 1.19 | 3.33 | 0.023 |
| miR-639 | 4.1 | 1.25 | 3.28 | 0.026 |
| miR-3125 | 4.1 | 0.81 | 5.09 | <0.001 |
| miR-3616-3p | 4.3 | 1.15 | 3.73 | 0.010 |
| miR-597-5p | 4.5 | 1.30 | 3.47 | 0.016 |
| miR-22-3p | 4.5 | 1.28 | 3.54 | 0.014 |
| miR-378f | 4.9 | 1.51 | 3.25 | 0.027 |

| *sncRNA* | *log$_2$ FC* | *SE* | *Wald statistic* | *Adj. p-values* |
|---|---|---|---|---|
| miR-1236-3p | 5.0 | 1.10 | 4.55 | <0.001 |
| miR-3658 | 5.1 | 1.42 | 3.58 | 0.014 |
| miR-1290 | 5.3 | 1.49 | 3.55 | 0.014 |
| miR-3646 | 5.7 | 1.89 | 3.02 | 0.048 |
| miR-6775-3p | 5.9 | 1.32 | 4.47 | <0.001 |
| miR-6132 | 6.7 | 1.86 | 3.57 | 0.014 |
| ***Lesser-expression*** | | | | |
| miR-4787-5p | -24.2 | 2.04 | -11.86 | <0.001 |
| miR-188-3p | -24.0 | 2.43 | -9.91 | <0.001 |
| miR-944 | -23.9 | 2.21 | -10.80 | <0.001 |
| miR-7153-3p | -23.8 | 2.43 | -9.80 | <0.001 |
| miR-548g-5p | -8.1 | 2.03 | -4.01 | 0.004 |
| miR-3977 | -5.3 | 1.75 | -3.00 | 0.049 |
| miR-885-3p | -5.1 | 1.54 | -3.28 | 0.026 |
| miR-942-5p | -5.0 | 1.59 | -3.15 | 0.037 |
| miR-4667-5p | -4.4 | 1.28 | -3.41 | 0.019 |
| miR-93-5p | -1.5 | 0.36 | -4.21 | 0.002 |
| miR-486-5p | -1.2 | 0.29 | -4.04 | 0.004 |
| miR-1178-3p | -1.0 | 0.34 | -3.02 | 0.048 |
| miR-16-5p | -0.9 | 0.25 | -3.53 | 0.014 |
| piR_000765/gb/DQ570956/Homo | -2.3 | 0.74 | -3.13 | 0.039 |

*Abbreviations: sncRNA, small non-coding RNA; log2 FC, log2 fold change; SE, standard error associated with the LFC estimate; Adj. p-values, p-values adjusted for false discovery rate (FDR). Differentially expressed small non-coding RNAs (sncRNAs) between non-exposed (n=27) and exposed (n=39) participants. reviations:*

### *Supplementary Material Table S2:*

*Studies focusing on miRNA expression in neuroinflammation and neurodegenerative diseases or other neuropathological processes and exposure to metals and toxicants, for three prioritized miRNAs differentially expressed in exposed participants and correlated with whole-blood metal concentrations.*

| miRNA | Exposure to metals or other toxicants | Inflammation and other pathological processes (*in vitro* studies) | In PD | In ADRD (Alzheimer's disease and related disorders) | Other neurological disorders |
|---|---|---|---|---|---|
| miR-16-5p | ↓ workers exposed to Al and Sb[1]<br>↓ workers exposed to Hg[2]<br>↑ animal model exposed to Cd[3]<br>↑ *in vitro* treated with Mn[4]<br>↑ animals exposed to As[5]<br>↑animals co-exposed to Se+Pb[6] | ↓ Meningitic *E. coli* infection in human brain cells[7]<br>↓Aβ-treated cells[8]<br>↑ *in vitro* treated with TNF-α[9]<br>↑ PD cell model exposed to Mn[4] | ↑ *In vitro*[4]<br>↓ Serum[10] | ↓ CSF EV[11]<br>↓ CSF[12]<br>↑ CTX[13]<br>↓ *In vitro*[8,14]<br>↑ *In vitro*[15]<br>↑ Blood[16]<br>↓ Severity of dementia[17]<br>↓ YOAD[11] | ↓ Blood; CJD[16]<br>↑ CTX; CJD[18,19]<br>↓ Blood; ALS[20]<br>↑ MSA[21]<br>↑ *In vivo*; Prion disorder[22]<br>Prion disorder[23]<br>Cognitive impairment induced by mixed heavy metals[24]<br>Depression induced by mixed heavy metals[25] |
| miR-93-5p | ↓ workers exposed to solvents[26]<br>↓ women exposed to Pb and Hg[27] | ↓ Meningitic *E. coli* infection in human brain cells[7] | | ↑ Plasma EV[28]<br>↑ Serum EV[29]<br>↓ Serum[30]<br>↑ Blood[16]<br>↓ CTX[31] | ↑ MSA[21]<br>↓ Blood; CJD[16]<br>↓ Blood; ALS[32] |
| miR-486-5p | ↑ female workers exposed to Hg[2]<br>↓ Pb exposure[33] | ↓ promotes apoptosis in cerebral ischemia[34] | ↑ plasma[35]<br>↑ Colonic biopsies in PD[36] | ↓ CSF (YOAD)[11]<br>↓ plasma AD[35]<br>↑ Plasma AD[37] | ↑ CTX and ST; HD[38]<br>↑ Plasma; HD[39]<br>↑ Plasma; VaD[40] |

*Abbreviations: PD, Parkinson's disease; ADRD, Alzheimer's disease and related dementias; MSA, multiple system atrophy; ALS, amyotrophic lateral sclerosis; CJD, Creutzfeldt–Jakob disease; YOAD, young-onset Alzheimer's disease; HD, Huntington's disease; VaD, vascular dementia; EV, extracellular vesicles; CSF, cerebrospinal fluid; CTX, cortex; ST, striatum; E. coli, Escherichia coli; Al, aluminum; Hg, mercury; Cd, cadmium; Sb, antimony; Pb, lead; Se, selenium; Mn, manganese; As, arsenic. ↓, decreased miRNA expression; ↑, increased miRNA expression.*

#### ***Supplementary Material Table S3:***

*Pearson partial correlations between circulating neuron-derived miRNAs and R2* MRI measures across brain regions, adjusted for age. Values represent correlation coefficients (r) with corresponding p-values.*

| **Brain ROIs** | **miR-16-5p r (p-value)** | **miR-93-5p r (p-value)** |
|---|---|---|
| *RN* | **−0.47 (<0.001)** | **−0.37 (0.007)** |
| *DN* | 0.16 (0.243) | 0.20 (0.150) |
| *SNr* | 0.15 (0.278) | 0.19 (0.177) |
| *SNc* | −0.07 (0.636) | −0.08 (0.582) |
| *CN* | −0.02 (0.913) | −0.13 (0.356) |
| *GP* | 0.21 (0.180) | 0.11 (0.847) |
| *PUT* | −0.05 (0.731) | −0.14 (0.376) |

*Abbreviations: ROIs, regions of interest; RN, red nucleus; DN, dentate nucleus; SNr, substantia nigra pars reticulata; SNc, substantia nigra pars compacta; CN, caudate nucleus; GP, globus pallidus; PUT, putamen.*

#### ***Supplementary Material Table S4:***

*Mediation analysis between whole-blood lead (Pb) levels and R2* MRI signal in the red nucleus (RN), with independently miR-16-5p and miR-93-5p as mediator. Models were adjusted for age and subsequently for age and exposure group.*

| **Mediator** | **Adj.** | **Path a (Pb → miRNA) β (p)** | **Path b (miRNA → R2*) β (p)** | **Path c (direct effect) β (p)** | **Indirect effect β (95% CI)** | **Total effect β (p)** |
|---|---|---|---|---|---|---|
| **miR-16-5p** | ***Age*** | **−0.74 (<0.001)** | **−1.64 (0.007)** | **0.71 (0.40)** | **1.21 (0.36–2.47)** | **1.93 (0.015)** |
| **miR-16-5p** | ***Age + group*** | **−0.53 (0.012)** | **−1.59 (0.011)** | **0.60 (0.52)** | **0.85 (0.14–2.15)** | **1.45 (0.12)** |
| **miR-93-5p** | *Age* | **−0.92 (<0.001)** | −0.85 (0.088) | 1.15 (0.19) | 0.78 (−0.01–1.78) | 1.93 (0.015) |
| **miR-93-5p** | *Age + group* | **−0.57 (0.026)** | −0.78 (0.14) | 1.01 (0.30) | 0.45 (−0.06–1.32) | 1.45 (0.12) |

***Abbreviations:*** *β, regression coefficient p, p-values;* ***path a****, effect of blood Pb levels on miRNA expression;* ***path b****, effect of miRNA expression on R2* in the red nucleus;* ***path c****, direct association between Pb exposure and R2* RN after inclusion of the mediator in the model.*

#### *SUPPLEMENTARY MATERIAL FIGURE S1:*

*Deep-sequencing analysis of the serum-derived EV miRNAs from exposed and non-exposed. A) Volcano plot: The y-axis values show the negative logarithm base 10 (log10) of the p-value; the horizontal gray dashed line on the plot represents the threshold p-value set at 0.05. The x-axis values indicate the log2-fold change; the vertical red dashed lines represent the fold change threshold set to 2. The dots on the positive side represent greater-expressed miRNA, whereas the dots within the negative area represent lower-expressed miRNA. The significantly differentially expressed miRNAs are marked with blue dots, whereas red represents the non-significantly expressed miRNAs. B) MA plot: Each dot represents one miRNA, with each triangle referring to the value beyond the y-axis range. The color of the dots represents significance α = 0.05 one-tailed used for explorative research; blue means significant, whereas grey means not significant. The x-axis represents the number of counts for the miRNA, and the y-axis corresponds to the log2-fold change for the miRNA. The dot at the top half (positive) represents that this miRNA is greater-expressed, and the dot at the bottom half (negative) represents that this miRNA is lower-expressed in the exposed compared to non-exposed participants.*

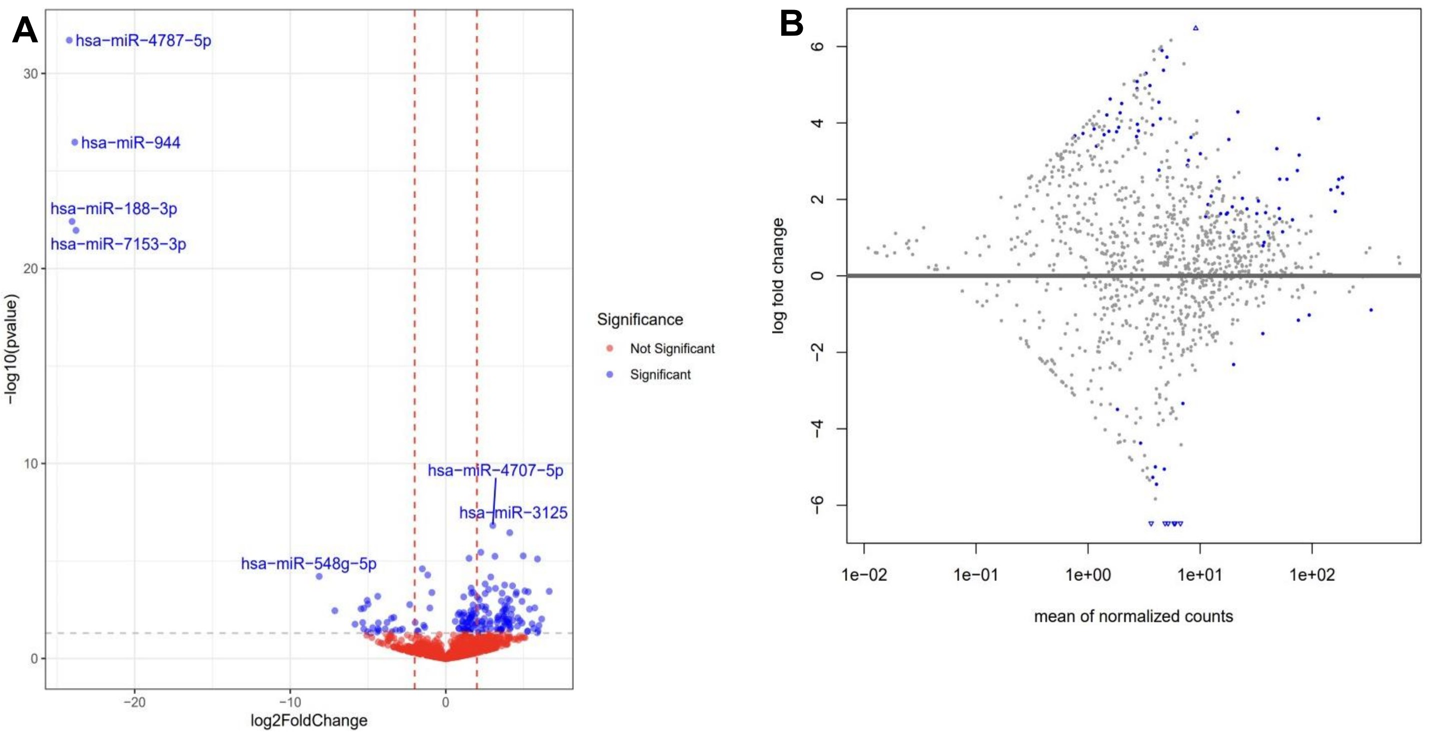

### *SUPPLEMENTARY MATERIAL FIGURE S2:*

*Deep-sequencing analysis of the 50 significantly deregulated miRNAs and 1 piRNA n exposed versus non-exposed participants.. A) Hierarchical clustering heatmap: Two-dimensional grid matrix derived from the functional heatmap in R displays the 50 differentially expressed miRNAs and 1 piRNA with the adj. p-value < 0.05 (rows) for the non-exposed (blue) and exposed participants (red) (columns). Each entry of the grid represents the mean (log2) fold change of the expression of a given miRNA on each sample. A blue grid corresponds to a low miRNA expression level (downregulation), whereas red represents a high miRNA expression level (upregulation), as indicated by the color key. B) PCoA plot: Investigates the resemblance of miRNA from non-exposed (black) and exposed (red). The lines of ellipses show 95% confidence intervals. Samples outside the ellipse line mean miRNA expression differ greatly from the rest of the samples. The p-value for the variance is the result of the permutation test, and the p-value for the mean is the result of the PERMANOVA test. C) LEfSe plot: Calculates the log LDA score for each miRNA labeled to the left. Grey bars represent miRNAs from the serum-derived EVs of non-exposed, and the blue bars represent those from exposed participants.*

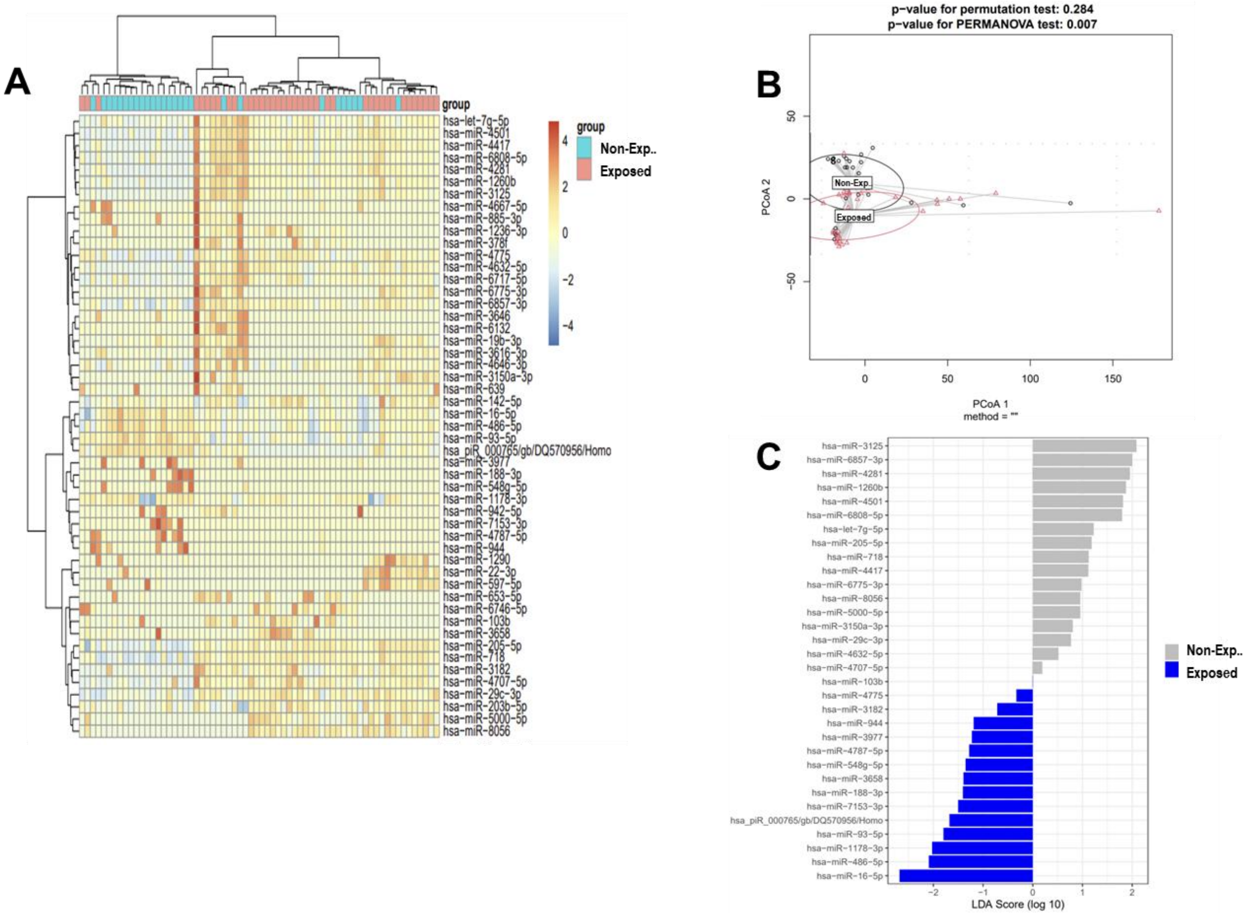

***SUPPLEMENTARY MATERIAL FIGURE S3:***

*Associations between whole-blood metal concentrations and miRNA expression in 66 participants (27 non-exposed, gray; 39 exposed participants, red). Scatterplots display whole-blood metal concentrations (natural log–transformed; Pb, Cu, Se, K; x-axis) versus miRNA expression (natural log–transformed; y-axis) for three miRNAs (miR-16-5p, miR-486-5p, and miR-93-5p). The black line represents the linear trend across all participants, with shaded areas indicating 95% confidence intervals.*

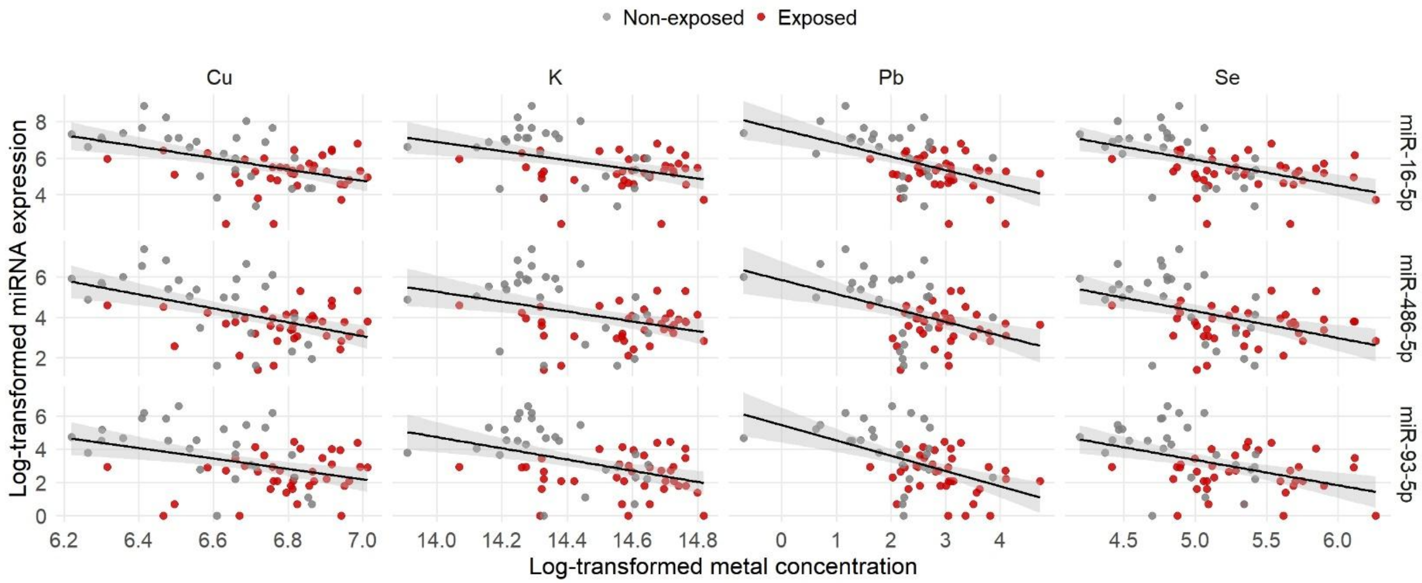

*Abbreviations: Cu, copper; K, potassium; Pb, lead; Se, selenium.*

### *Supplementary Material Figure S4:*

*Associations between welding exposure metrics and log-transformed miRNA expression in 66 participants (27 non-exposed, gray; 39 exposed participants, red). Scatterplots display welding exposure metrics (ELT, YrsW, E90, HrsW; x-axis) versus miRNA expression (natural log–transformed; y-axis) for three miRNAs (miR-486-5p, miR-16-5p, and miR-93-5p). The black line represents the linear trend across all participants, with shaded areas indicating 95% confidence intervals.*

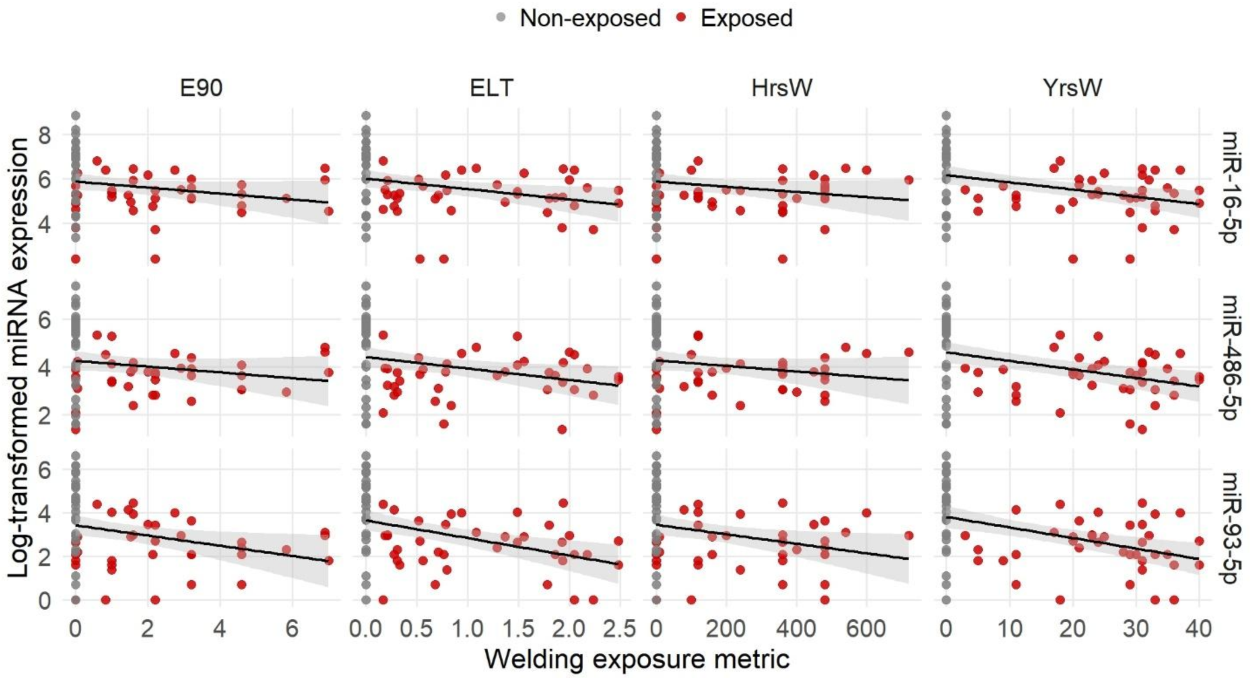

*Abbreviations: YrsW, years welding; ELT, lifetime cumulative welding exposure; E90, cumulative 90-day welding exposure; HrsW, hours welding*

### ***SUPPLEMENTARY MATERIAL FIGURE S5:***

*Overall mixture effect estimates, adjusted for age(left) and subsequently for age and exposure group (right) on miR-16-5p (A-B), miR-93-5p (C-D), and miR-486-5p expression (E-F). The x-axis represents quantiles of the joint metals distribution, with the 50th percentile (q = 0.50) as the reference level. Points indicate mean differences of the overall mixture in miRNA expression relative to the 50th percentile level, and vertical bars represent 95% credible intervals (CrI).*

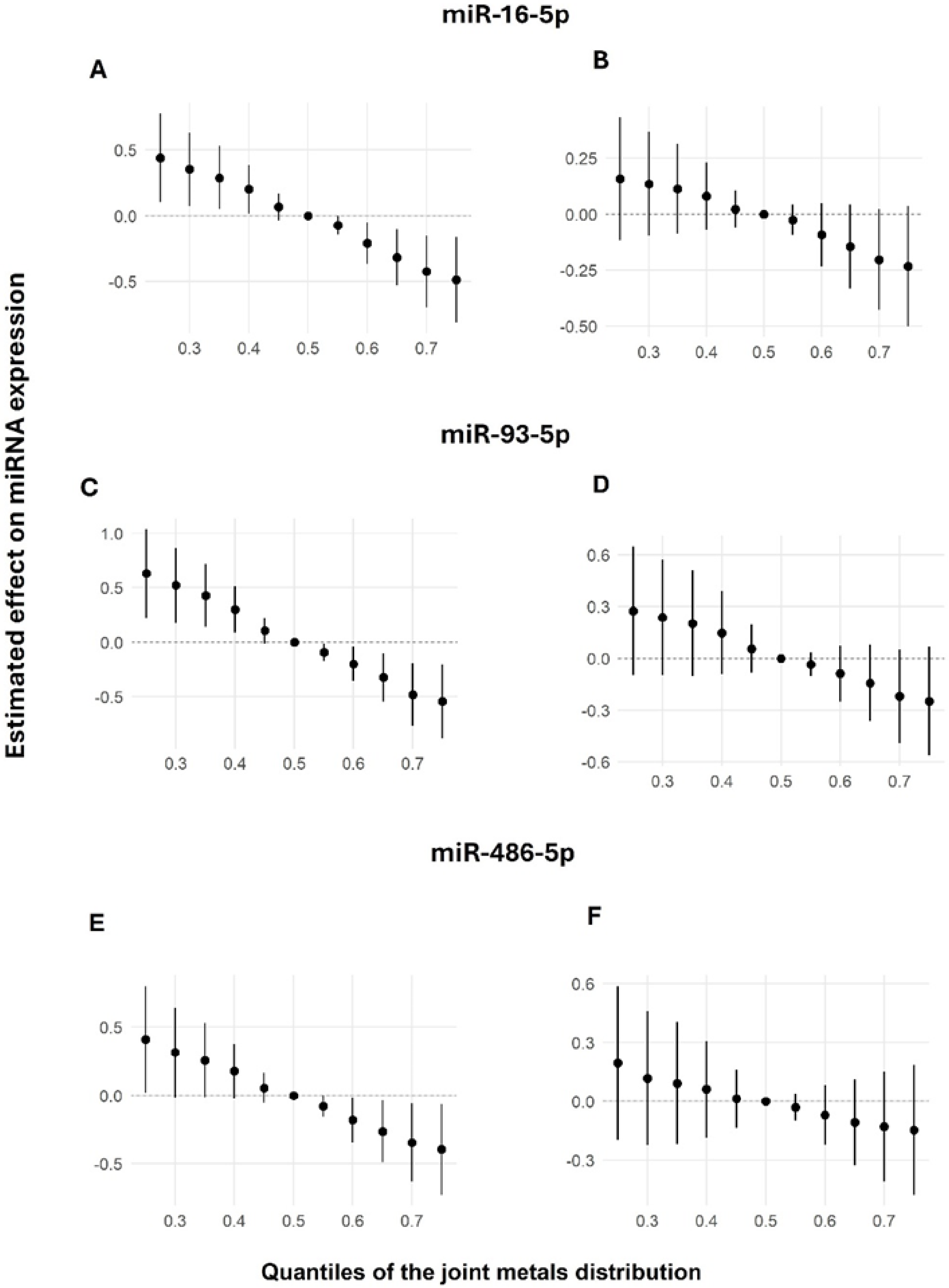

***SUPPLEMENTARY MATERIAL FIGURE S6:***

*Single-metal exposure–response between individual log-transformed metal levels in whole blood and log-transformed expression levels of: A) miR-16-5p, B) miR-93-5p, and C) miR-486-5p. Models adjusted for age and exposure groups. Blue lines indicate mean estimated effects, and shaded areas represent the 95% credible intervals, holding all other metals at their 50th percentile. Metals included: copper (Cu), iron (Fe), lead (Pb), magnesium (Mg), manganese (Mn), potassium (K), selenium (Se), sodium (Na), strontium (Sr), and zinc (Zn).*

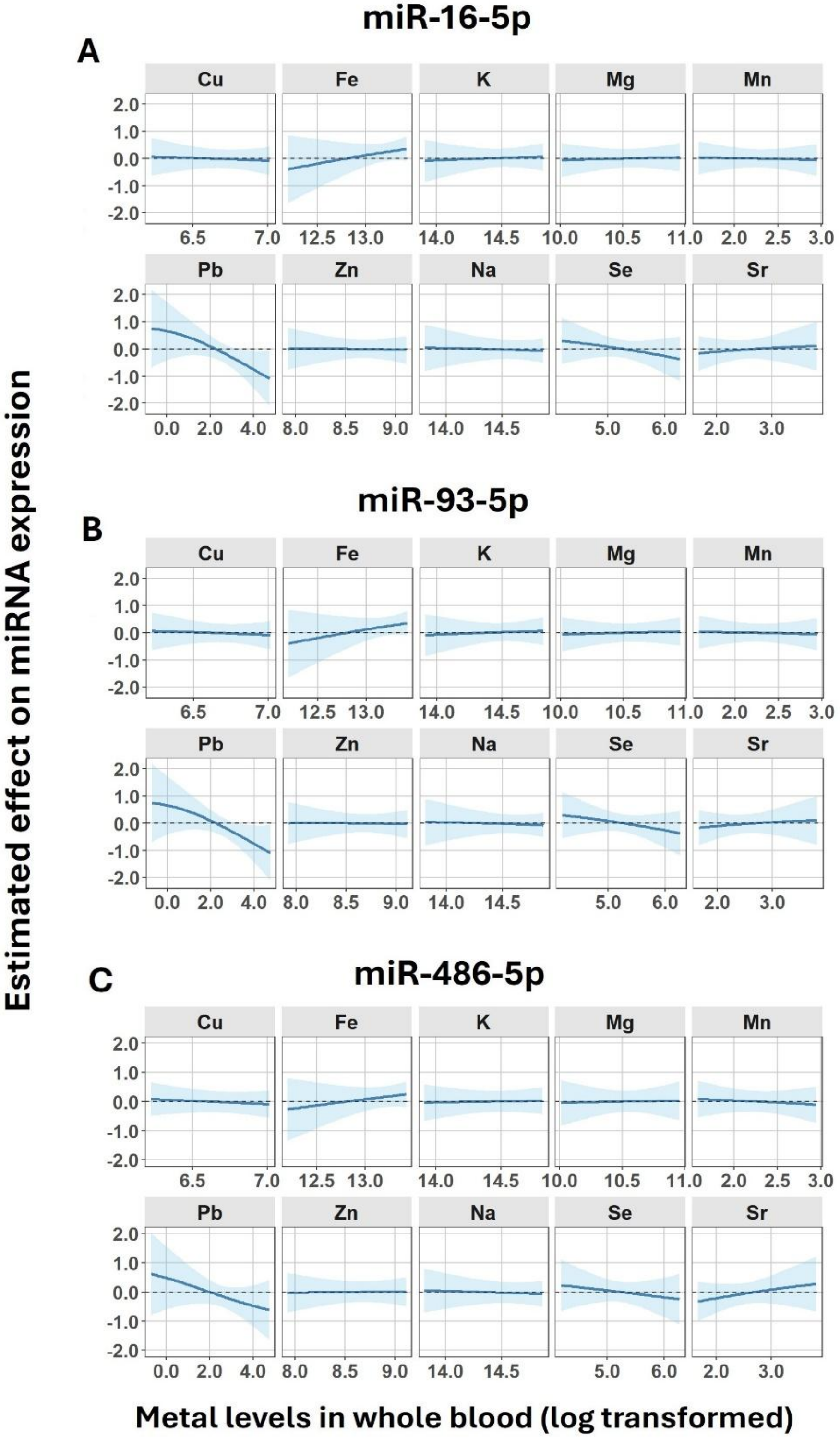

***Supplementary Material Figure S7:***

*Bivariate exposure–response between individual log-transformed metal levels in whole blood (x-axis; expo 1) and log-transformed expression levels of miR-16-5p conditional on another metal (expo 2) held at the 25th (red), 50th (green), or 75th (blue) percentiles. Models adjusted for age and exposure groups. Metals included: copper (Cu), iron (Fe), lead (Pb), magnesium (Mg), manganese (Mn), potassium (K), selenium (Se), sodium (Na), strontium (Sr), and zinc (Zn).*

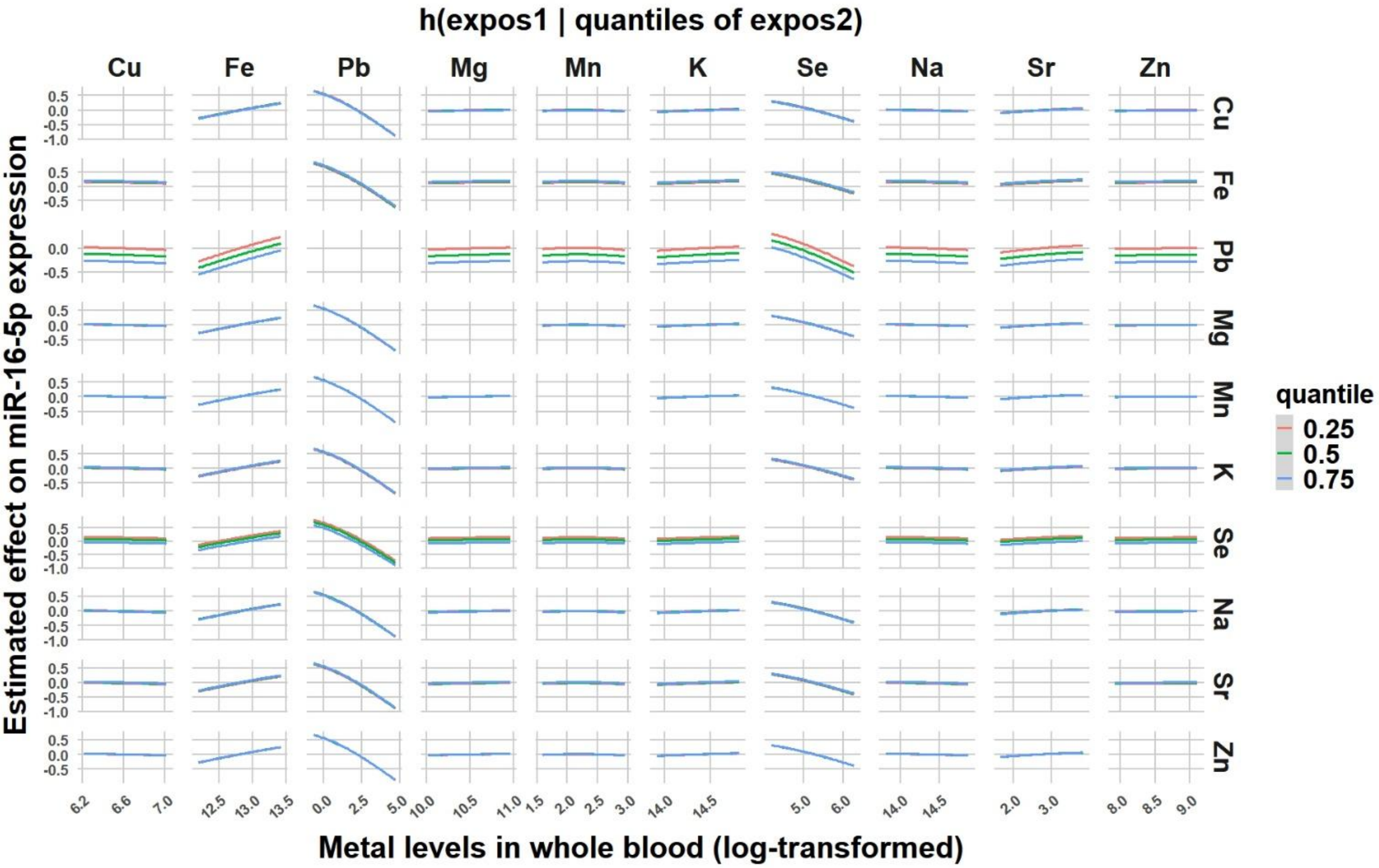

***SUPPLEMENTARY MATERIAL FIGURE S8:***

*Bivariate exposure–response between individual log-transformed metal levels in whole blood (x-axis; expo 1) and log-transformed expression levels of miR-93-5p conditional on another metal (expo 2) held at the 25th (red), 50th (green), or 75th (blue) percentiles. Models adjusted for age and exposure groups. Metals included: copper (Cu), iron (Fe), lead (Pb), magnesium (Mg), manganese (Mn), potassium (K), selenium (Se), sodium (Na), strontium (Sr), and zinc (Zn).*

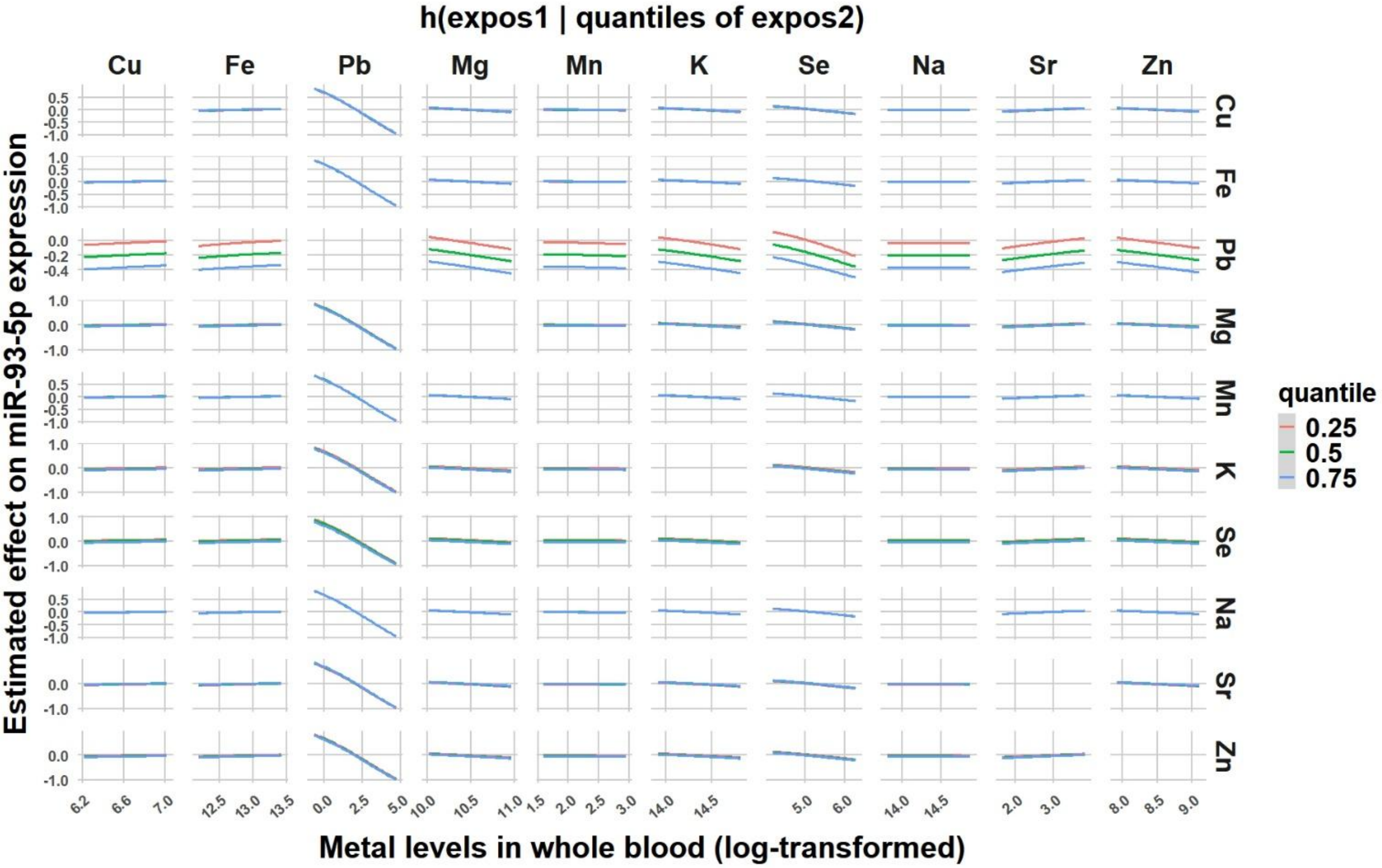

***Supplementary Material Figure S9:***

*Bivariate exposure–response between individual log-transformed metal levels in whole blood (x-axis; expo 1) and log-transformed expression levels of miR-486-5p conditional on another metal (expo 2) held at the 25th (red), 50th (green), or 75th (blue) percentiles. Models adjusted for age and exposure groups. Metals included: copper (Cu), iron (Fe), lead (Pb), magnesium (Mg), manganese (Mn), potassium (K), selenium (Se), sodium (Na), strontium (Sr), and zinc (Zn).*

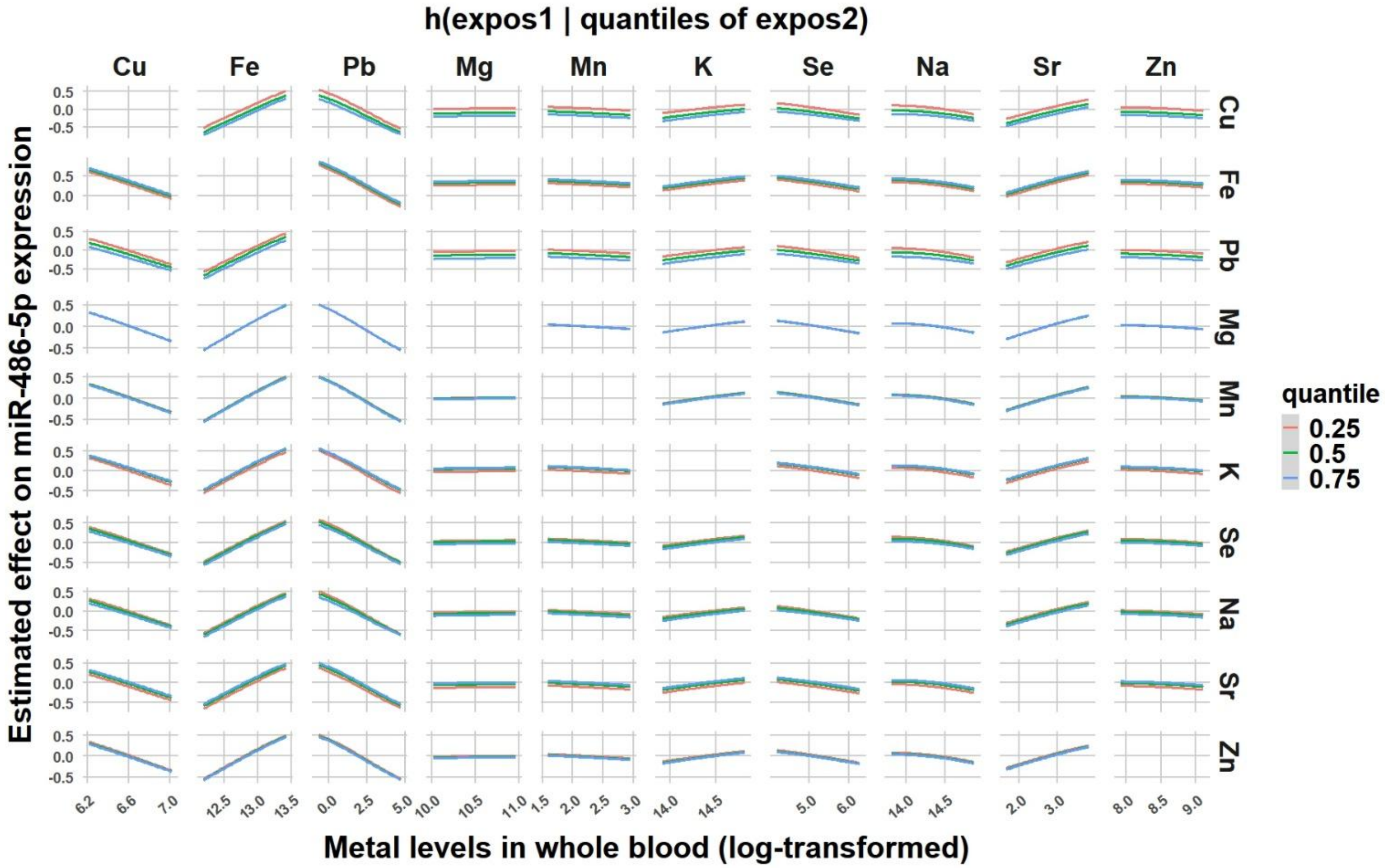

### ***SUPPLEMENTARY MATERIAL FIGURE S10:***

*Single-variable risk for each metal on expression levels of miR-16-5p (left), miR-93-5p (center), and miR-486-5p (right), conditional on other metals held at the 25th, 50th and75th percentile (black, blue, and red, respectively). Models adjusted for age and exposure groups.*

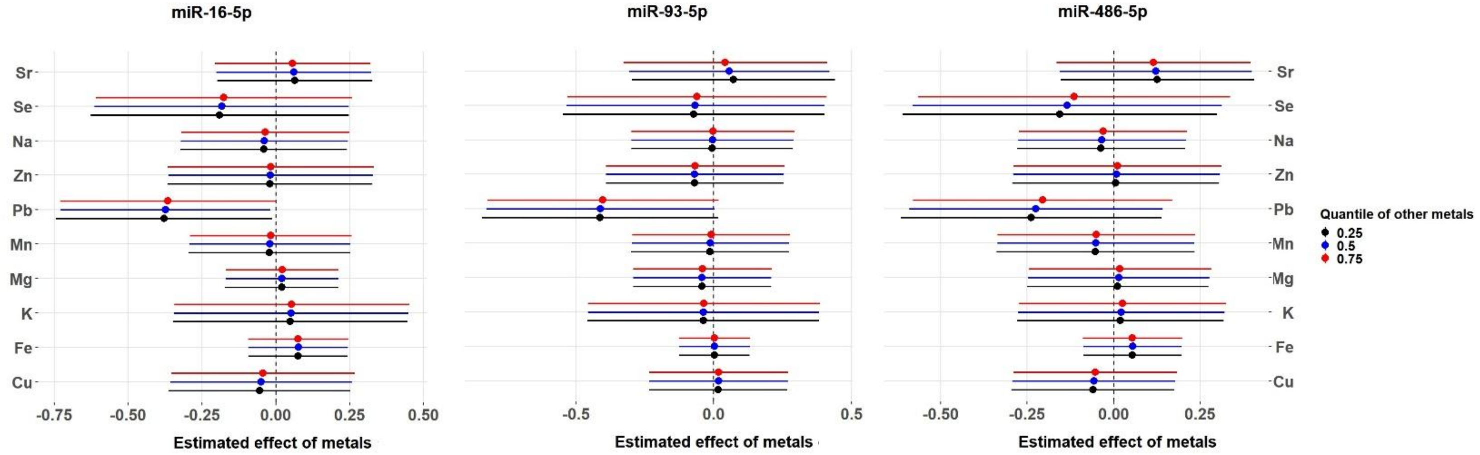
